# Shared risk factors for malaria and schistosomiasis co-infection: a systematic review and meta-analysis

**DOI:** 10.64898/2025.12.10.25341957

**Authors:** Max M. Lang, Bethany Lyne, Christl A. Donnelly, Goylette F. Chami

## Abstract

**Background:** Malaria and schistosomiasis are co-endemic across sub-Saharan Africa, resulting in frequent co-infection, yet the shared risk factors driving co-infection remain poorly synthesized.

**Methods:** We conducted a systematic review and meta-analysis to identify shared risk factors for malaria-*Schistosoma* co-infection and to narratively synthesize the statistical methodologies applied in the literature. We searched PubMed/MEDLINE, Embase, Web of Science, Global Index Medicus, and Global Health from inception to February 19, 2025 (PROSPERO CRD420250648824). We pooled effect sizes for risk factors across sociodemographic, environmental, and behavioral dimensions. Fixed-effects meta-analysis with inverse variance weighting was used to calculate pooled Odds Ratios (OR) and 95% confidence intervals (CIs). Study quality was assessed using a modified version of the Quality Assessment tool for Observational Cohort and Cross-Sectional Studies by the National Institutes of Health.

**Results:** We screened 1,345 records and included 30 studies conducted across 12 African countries. A meta-analysis of 27 studies showed that schistosomiasis infection was associated with 1.27 times higher odds of malaria (OR 1.27; 95% CI: 1.17–1.39). Narrative synthesis identified age as an important predictor, with risk consistently peaking in older children and adolescents (typically 8–17 years). Sex influences were setting-dependent: males had significantly higher odds of co-infection in community-based studies (OR 2.08; 95% CI: 1.64–2.63), whereas no significant association was found in school-based studies (OR 0.87; 95% CI: 0.64–1.19). Direct water contact was strongly associated with co-infection (OR 2.53; 95% CI: 1.60–4.00). Heterogeneity was high (*I*^2^ *>* 80%). Only one study was categorized as high risk of bias.

**Conclusion:** The association between malaria and schistosomiasis appears driven by overlapping environmental and behavioral exposures, specifically water contact in older children.

**Author summary:** Malaria and schistosomiasis are parasitic diseases that frequently infect the same individuals, particularly in sub-Saharan Africa. While it is known that co-infection exacerbates clinical outcomes like anemia, the specific behaviors and environmental conditions that put individuals at risk of co-infection are not well understood. We performed a systematic review of 30 studies to identify risk factors for co-infection. We found that individuals infected with schistosomiasis were 27% more likely to harbor malaria. Our analysis suggests this relationship is largely driven by shared risk factors rather than biological interaction. Risk appears to peak in older children and adolescents (ages 8–17 years). Gender roles influence risk differently depending on the setting: males were at higher risk in community settings, likely due to occupational activities like fishing or farming, but this risk disappeared in school settings where boys and girls have similar daily routines. Direct water contact was a risk factor for co-infection, but it was inconsistently measured across studies. Our findings support the case for investigation of interventions to support integrated disease control for schistosomiasis and malaria.

## Introduction

Malaria and schistosomiasis are among the most prevalent parasitic diseases in low- and middle-income countries. Globally, malaria accounted for an estimated 263 million cases, while schistosomiasis affected approximately 240 million people [1, 2]. The burden of both infections is heavily concentrated in sub-Saharan Africa, creating a significant geographic overlap of at-risk populations [2, 1, 3, 4]. Individual co-infection with both *Plasmodium* and *Schistosoma* parasites is common [3], and its influence on clinical outcomes is well-established [5, 6, 7, 8]. Despite the clinical importance, the risk factors for acquiring co-infections remain poorly understood.

Individual empirical studies have investigated a range of potential risk factors for co-infection, with sociodemographic factors like age and sex being the most frequently studied. There is agreement, for example, that younger age is a risk factor for co-infection [9, 10], though there is disagreement on what age range [11, 12, 13]. Conversely, findings for sex are more equivocal; males are frequently reported at higher risk, but this association is often tied to behavioral patterns, such as occupation or recreational water contact, and is not a universal finding [3, 14]. Factors related to behavior and environment, such as proximity to water bodies or specific water contact activities, are also commonly linked to co-infection, though their relative contributions vary [3, 10]. Other factors, including socioeconomic status or the use of preventative measures like bednets, have been less frequently assessed [9, 4]. Thus, while some consensus exists, the magnitude of these effects and the reasons for their inconsistency across different settings remain poorly synthesized.

Existing systematic reviews have provided some insight for co-infection, but their focus has been primarily on estimating pooled prevalence or evaluating clinical outcomes rather than synthesizing risk factors. For instance, past reviews have focused on specific populations, such as children [15], or specific regions, like Nigeria [16], to quantify the burden of co-infection. Afolabi et al. [15] found a pooled malaria-helminth co-infection prevalence of 17.7% in children and noted that the impact on outcomes like anemia remained inconclusive. Similarly, Ojo et al. [16] estimated a 15% co-infection prevalence in Nigeria, highlighting a lack of well-designed studies.

Beyond empirical reviews on schistosomiasis and malaria co-infection, there have been methodological reviews. A methodological review by Powell-Romero et al. [17] evaluated statistical approaches to modeling co-infections, while Duguay et al. [3] investigated the integration of prevention and control programs. Further, a recent meta-analysis by Koellsch-Amet et al. [18] confirmed that malaria-schistosome co-infection occurs more frequently than would be expected by chance. To date, however, no review has systematically synthesized the individual-level risk factors associated with co-infection.

We performed a systematic review and meta-analysis to synthesize the available evidence and address the following questions: 1) What are the risk factors associated with co-infection of malaria and schistosomes in humans? 2) What statistical assumptions have been made about the relationship between these infections in the literature?.

## Materials and methods

### Protocol and search strategy

The protocol for this systematic review was first submitted and registered with PROSPERO on February 18, 2025 and February 24, 2025 (CRD420250648824)[19], respectively. The review is reported in accordance with the Preferred Reporting Items for Systematic Reviews and Meta-Analyses (PRISMA) 2021 guidelines [20].

We searched six electronic databases from inception to February 19, 2025. Databases searched included PubMed/MEDLINE (1946-), Embase via Ovid (1974-), Web of Science (1964-), Global Index Medicus (1901-), and Global Health (1973-). The search strategy combined Medical Subject Heading (MeSH) terms and free text using the following string: (malaria* OR Plasmodium OR Plasmodium falciparum OR ”P. falciparum” OR ”Plasmodium vivax”) AND (schistosom* OR bilharzia* OR Schistosoma mansoni OR Schistosoma haematobi* OR helminthiasis OR ”snail fever”) AND (co-infection* OR ”dual infection” OR ”co-occurrence” OR ”co-occur” OR ”co-distributed” OR ”co-distribution” OR cooccur* OR codistribut* OR ”concomitant” OR ”polyparasitism” OR ”multiparasitism” OR ”multi-infection” OR ”simultaneous infection” OR ”joint infection” OR ”superinfection” OR ”synergistic infection”). Results were filtered for studies on humans, with any remaining animal studies excluded during screening. The expanded search strings and database results can be found in Supplementary Text S1.

### Inclusion and exclusion criteria

Eligible study designs included cohort studies, cross-sectional studies, case-control studies, before-after studies (reporting baseline infection and exposure), and randomised controlled trials (reporting baseline and/or control group data). Studies were required to report odds ratios (OR), hazard ratios (HRs), relative risks (RRs), or sufficient information to reconstruct these effect sizes for individual-level associations between risk factors and schistosome-malaria co-infection. We excluded studies using self-reports or indirect approximations of infection status (e.g., diarrhoea for *S. mansoni*, fever for malaria, microhematuria for *S. haematobium*), simulation-only studies, mathematical models, reviews, meta-analyses, case reports, and editorial articles. Studies were required to have abstracts and titles in English, though no restrictions were placed on full-text language, with native speakers assisting with translation where needed.

### Screening and data extraction

Two reviewers (MML and BL) independently screened titles and abstracts using Covidence [21], with duplicates removed automatically (578 duplicates) and manually (86 duplicates). The full text of potentially eligible articles was screened independently by both reviewers. Disagreements were resolved by a third reviewer (GFC). Data extraction was performed by one reviewer (MML) using a pre-tested standardized form in Microsoft Excel (Version 16.102.1 (25101829)) that was adapted from Kmentt et al. [22]. The extraction was performed via a two-step process. First, all covariates reported were extracted and summarized into a list of exposures. Based on this list of covariates they were then synthesized into broader domains of risk factors, where appropriate, such as spatial factors (e.g., distance to healthcare facilities, proximity to water bodies), and behavioral domains (e.g., water contact), among others. A randomly selected 10% of full-text publications underwent verification by a second reviewer (BL). When studies contained duplicated data from the same population, i.e. same population and study years, the study with the larger sample size was retained. One attempt was made to contact authors for additional information when essential data were missing.

### Outcome and exposures

The primary outcome was co-infection with malaria (any *Plasmodium* species) and schistosomiasis (any *Schistosoma* species), with co-infection defined as the concurrent or sequential occurrence of both infections, regardless of temporal order, provided both were explicitly modeled as the dependent variable. We also investigated schistosomiasis infection as a risk factor for malaria. We extracted all covariates reported and categorized them into broader domains of potential risk factors for schistosome-malaria co-infection. Spatial factors (e.g., distance to healthcare facilities, proximity to water bodies, presence of water bodies) were grouped together, as well as any form of reported water contact (e.g., swimming, visiting backwater). The full data extraction table can be found in Supplementary Dataset S1.

### Subgroup analysis

Subgroup analyses were performed to explore sources of heterogeneity and effect modification when at least three studies reported on the same dimension of risk factor that could be synthesized. For the analysis of schistosomiasis as a risk factor for malaria infection, we conducted subgroup analyses by: *Schistosoma* species (*S. mansoni*, *S. haematobium*, or any schistosomiasis), study design (e.g., cross-sectional, cohort), Risk of bias (ROB) (low, moderate, high), reference population (uninfected M-S- vs. single-infection with malaria M+S-), effect estimate type (adjusted vs. unadjusted OR), and study setting (community, school, or health center). For the analysis of sex as a risk factor for co-infection, we stratified studies by study setting, ROB, reference population, and effect estimate type. A subgroup analysis by study design was not performed as only one study had a different design. Water contact as a risk factor was analyzed as a single pooled analysis without subgroup stratification, as only three studies were available.

### Risk of bias assessment

Study quality was assessed using a modified version of the Quality Assessment Tools for Observational Cohort and Cross-Sectional Studies from the National Institutes of Health (NIH), previously applied to schistosomiasis studies and adapted for malaria co-infection [23, 24]. Each criterion was scored as yes if met (1 point) or no/unclear if not met (0 points), yielding scores from 0-9. Studies were classified as low (score 7-9), moderate (4-6), or high (0-3) ROB. ROB assessments were primarily conducted by one reviewer (MML). A second reviewer (BL) independently cross-checked a 10% sample of the studies to ensure reliability, with any disagreements resolved by a third reviewer (GFC). When at least three studies per category were available, subgroup analyses were stratified by ROB. Otherwise, sensitivity analyses excluded high-risk studies to assess their influence on pooled estimates. The detailed ROB assessment can be found in Supplementary Dataset S2.

### Data analysis

All statistical analyses were performed using R version 4.2.1. Meta-analyses were implemented using the meta package (version 6.5-0). We first identified risk factors eligible for quantitative synthesis. A meta-analysis was performed for a specific exposure if at least three studies reported a comparable effect measure. We conducted three primary meta-analyses examining: sex as a risk factor for co-infection, schistosomiasis infection as a risk factor for malaria infection, and water exposure as a risk factor for co-infection. We prioritized adjusted odds ratios (aOR) when provided. If adjusted estimates were not available, we used the reported crude odds ratio (cOR). If no effect measure was reported but sufficient data were present (e.g., in a 2x2 table), we calculated the cOR and 95% CIs (see Supplementary Text S2). To avoid violating the assumption of independent estimates, each study was included only once in the main pooled meta-analysis. When studies presented multiple statistical models (e.g., for different subgroups or with different covariate sets), we extracted the effect measure from the model that was fitted to the entire study population. Species-specific estimates were used only in the species subgroup analysis.

For all pooled analyses, common-effects models were fitted using inverse variance weighting with the assumption that all studies estimate the same underlying effect size. Forest plots were generated displaying study-specific and pooled effect estimates, with studies weighted according to the inverse of their variance. Between-study heterogeneity was quantified using the *I*^2^-statistic. Funnel plots were constructed to assess publication bias visually; this inspection was supplemented by Egger’s test to formally assess for small-study effects, with *p <* 0.1 considered indicative of significant asymmetry [25].

For findings that could not be pooled (i.e., those reported by fewer than three studies or with different definitions, e.g., study sites, age groups), we performed a narrative synthesis by first summarizing the statistical methodologies, model setups, variable selection techniques, and covariates used across the included studies. Following this synthesis, we categorized the remaining risk factors into relevant domains (e.g., sociodemographic, behavioral, environmental) and summarized the reported associations, noting the direction of effect and illustrative effect sizes.

## Results

The search returned 1,345 records. After removing 664 duplicates (578 automatically by Covidence and 86 manually), 681 titles and abstracts were screened. Following this, 569 studies were excluded, and 112 full-text articles were assessed for eligibility, of which 82 studies were excluded. The primary reasons for exclusion were ineligible outcomes (41.5%, 34/82), being a descriptive study design with no associational analysis (23.2%, 19/82), or the full text being unavailable (11.0% 9/82). A total of 30 studies were included in the final systematic review (Figure 1). Of these 30 studies, representing 16,775 participants, 27 (90%) provided comparable outcome and exposure measures and were included in at least one quantitative meta-analysis, the remaining three studies (see Supplementary Dataset S1) were synthesized narratively.

**Fig 1.**
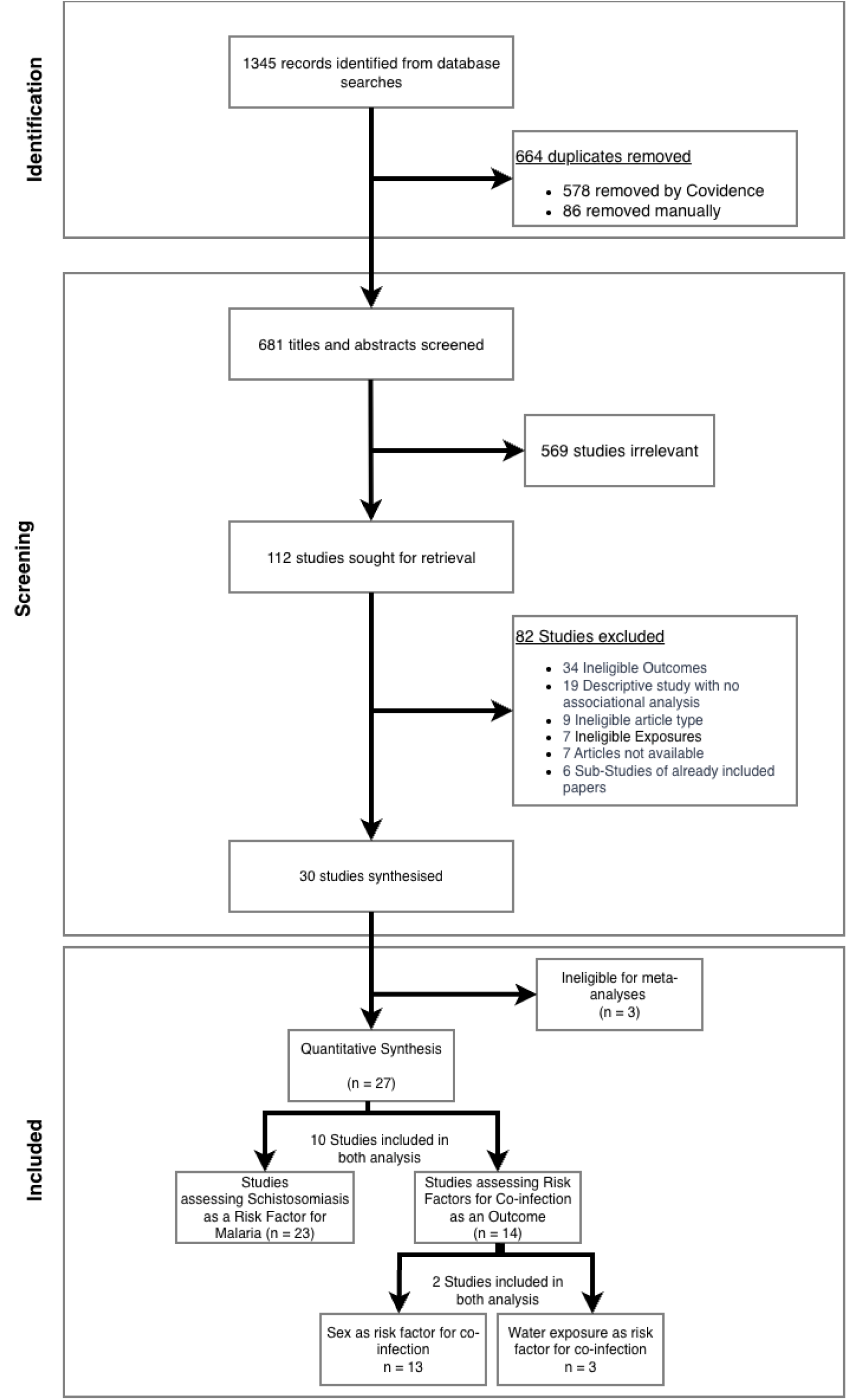
Study selection flowchart. PRISMA flowchart (Preferred Reporting Items for Systematic Reviews and Meta-Analyses) depicting the number of records identified, screened, assessed for eligibility, and included in the review, with reasons for exclusions at each stage.

### Study characteristics

Summary characteristics of all 30 included studies are reported in Table 1. All included studies were conducted in sub-Saharan Africa, across 12 different countries (Table 2). 27 studies reporting 30 effect sizes were included in the meta-analysis of the association between schistosomiasis infection and malaria. Ethiopia (16.7%, 5/30), Nigeria (13.3%, 4/30), and Senegal (13.3%, 4/30) were the most represented countries, accounting for over 40% of all included studies. The most frequent study design was cross-sectional (76.7%, 23/30), followed by cohort studies (20.0%, 6/30). Studies were predominantly conducted in rural localities (80.0%, 24/30) and school-based settings (36.7%, 11/30). For schistosome infection, studies most commonly reported on *S. haematobium* (66.7%, 20/30) or *S. mansoni* (60.0%, 18/30). The most common diagnostic methods for schistosomiasis were the Kato-Katz technique (46.7%, 14/30) and urine filtration (43.3%, 13/30). Malaria infection was most frequently diagnosed using microscopy (83.3%, 25/30), with 18 studies (60.0%) relying on it exclusively.

**Table 1.**
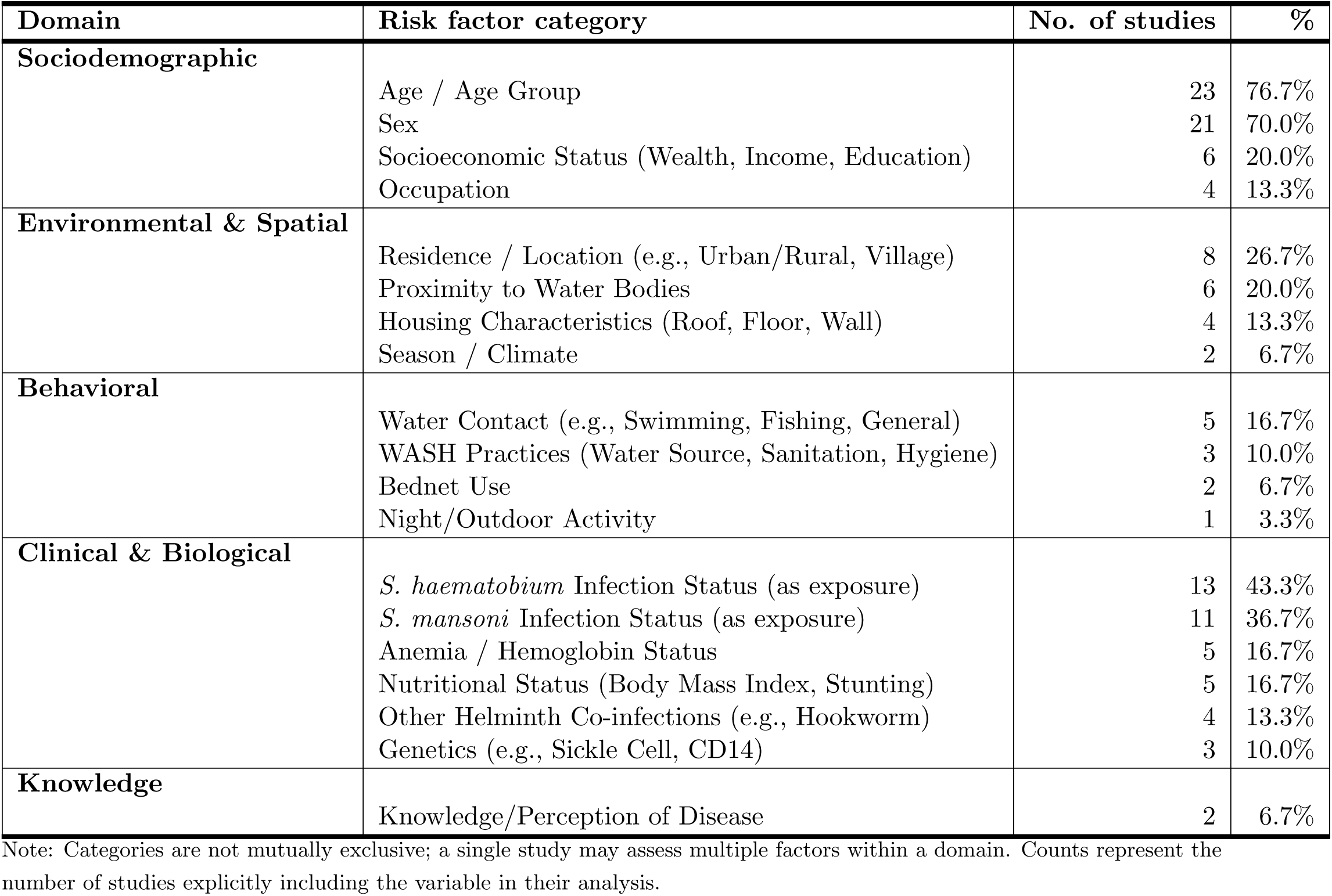
Frequency of Risk Factors and Exposures Assessed Across Included Studies (N = 30).

**Table 2.**
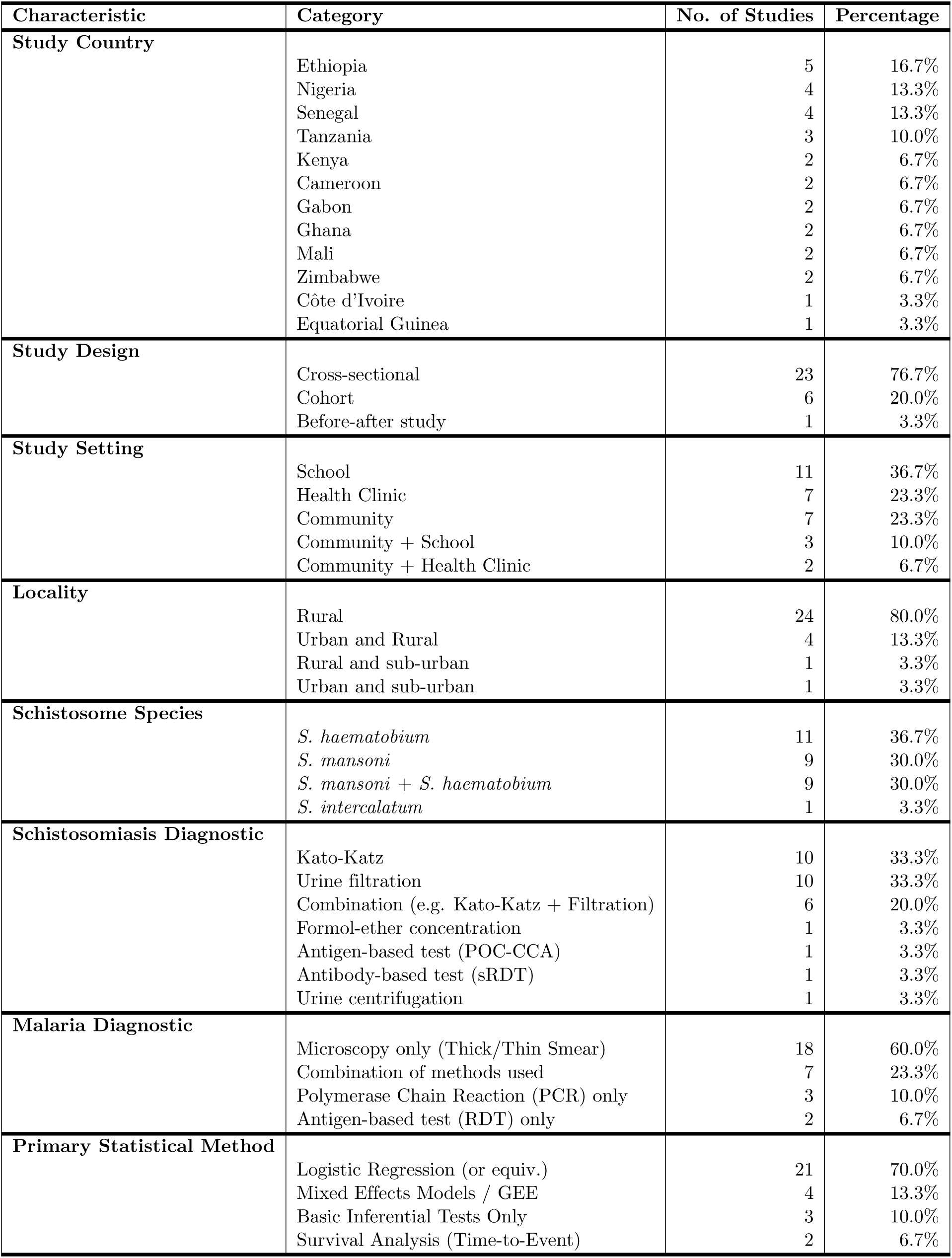
Summary of study characteristics (N = 30).

| Characteristic | Category | No. of Studies | Percentage |
| --- | --- | --- | --- |
| Study Country | Ethiopia | 5 | 16.7% |
|  | Nigeria | 4 | 13.3% |
|  | Senegal | 4 | 13.3% |
|  | Tanzania | 3 | 10.0% |
|  | Kenya | 2 | 6.7% |
|  | Cameroon | 2 | 6.7% |
|  | Gabon | 2 | 6.7% |
|  | Ghana | 2 | 6.7% |
|  | Mali | 2 | 6.7% |
|  | Zimbabwe | 2 | 6.7% |
|  | Côte d'Ivoire | 1 | 3.3% |
|  | Equatorial Guinea | 1 | 3.3% |
| Study Design | Cross-sectional | 23 | 76.7% |
|  | Cohort | 6 | 20.0% |
|  | Before-after study | 1 | 3.3% |
| Study Setting | School | 11 | 36.7% |
|  | Health Clinic | 7 | 23.3% |
|  | Community | 7 | 23.3% |
|  | Community + School | 3 | 10.0% |
|  | Community + Health Clinic | 2 | 6.7% |
| Locality | Rural | 24 | 80.0% |
|  | Urban and Rural | 4 | 13.3% |
|  | Rural and sub-urban | 1 | 3.3% |
|  | Urban and sub-urban | 1 | 3.3% |
| Schistosome Species | *S. haematobium* | 11 | 36.7% |
|  | *S. mansoni* | 9 | 30.0% |
|  | *S. mansoni* + *S. haematobium* | 9 | 30.0% |
|  | *S. intercalatum* | 1 | 3.3% |
| Schistosomiasis Diagnostic | Kato-Katz | 10 | 33.3% |
|  | Urine filtration | 10 | 33.3% |
|  | Combination (e.g. Kato-Katz + Filtration) | 6 | 20.0% |
|  | Formol-ether concentration | 1 | 3.3% |
|  | Antigen-based test (POC-CCA) | 1 | 3.3% |
|  | Antibody-based test (sRDT) | 1 | 3.3% |
|  | Urine centrifugation | 1 | 3.3% |
| Malaria Diagnostic | Microscopy only (Thick/Thin Smear) | 18 | 60.0% |
|  | Combination of methods used | 7 | 23.3% |
|  | Polymerase Chain Reaction (PCR) only | 3 | 10.0% |
|  | Antigen-based test (RDT) only | 2 | 6.7% |
| Primary Statistical Method | Logistic Regression (or equiv.) | 21 | 70.0% |
|  | Mixed Effects Models / GEE | 4 | 13.3% |
|  | Basic Inferential Tests Only | 3 | 10.0% |
|  | Survival Analysis (Time-to-Event) | 2 | 6.7% |

Studies assessed a wide range of potential risk factors, which were synthesized into key domains as summarized in Table 1. The most commonly assessed exposures were sociodemographic, particularly age (76.7%, 23/30) and sex (70.0%, 21/30). Current infection status was also frequently analyzed as an exposure for co-infection, with 13 studies (43.3%) assessing the role of *S. haematobium* infection and 11 studies (36.7%) assessing *S. mansoni* infection. Other domains, assessed less frequently, included environmental factors such as residence or proximity to water (26.7%, 8/30), behavioral factors like water contact (16.7%, 5/30) and Water, Sanitation, and Hygiene (WASH) related variables (10.0%, 3/30), and preventative measures such as bednet use (6.7%, 2/30), as detailed in Table 1.

### Risk of bias assessment

The overall quality of the 30 included studies was assessed as low-to-moderate ROB (see Supplementary Dataset S2). Based on our 9-point criteria, 10 studies (33.3%) were classified as having a low ROB (score 7-9), 19 studies (63.3%) as moderate risk (score 4-6), and one study (3.3%) as high risk (score 0-3). The classification of studies as moderate or high risk was primarily driven by lack of independent or blinded outcome assessment (80.0%, 24/30), failure to report participation rates (63.3%, 19/30), and ambiguous definitions of co-infection (60.0%, 18/30) (see Supplementary Table S2).

### Association of schistosome infection with malaria

Schistosome infection was associated with a 1.27 times increase in the odds of malaria infection (95% CI: 1.17–1.39; 27 studies; Figure 3). There was substantial and significant heterogeneity across the included studies (*I*^2^ = 86.4%, *p <* 0.0001). Among the 11 studies reporting specifically on *S. haematobium*, six (54.5%) identified a statistically significant positive association. In contrast, of the nine studies reporting on *S. mansoni*, only one reported a significant positive association. The remaining seven studies fell into the ’Schistosomiasis (Any)’ category, referring to studies where authors reported infection as a composite variable (positive for either *S. mansoni* or *S. haematobium*) without providing species-specific estimates; it did not imply concurrent co-infection with both schistosome species. Visual inspection of the funnel plot (Supplementary Figure S1) did not suggest asymmetry. This was confirmed by Egger’s regression test, which indicated no evidence of significant small-study effects (*t* = *−*0.41*, df* = 21*, p* = 0.6885).

**Fig 2.**
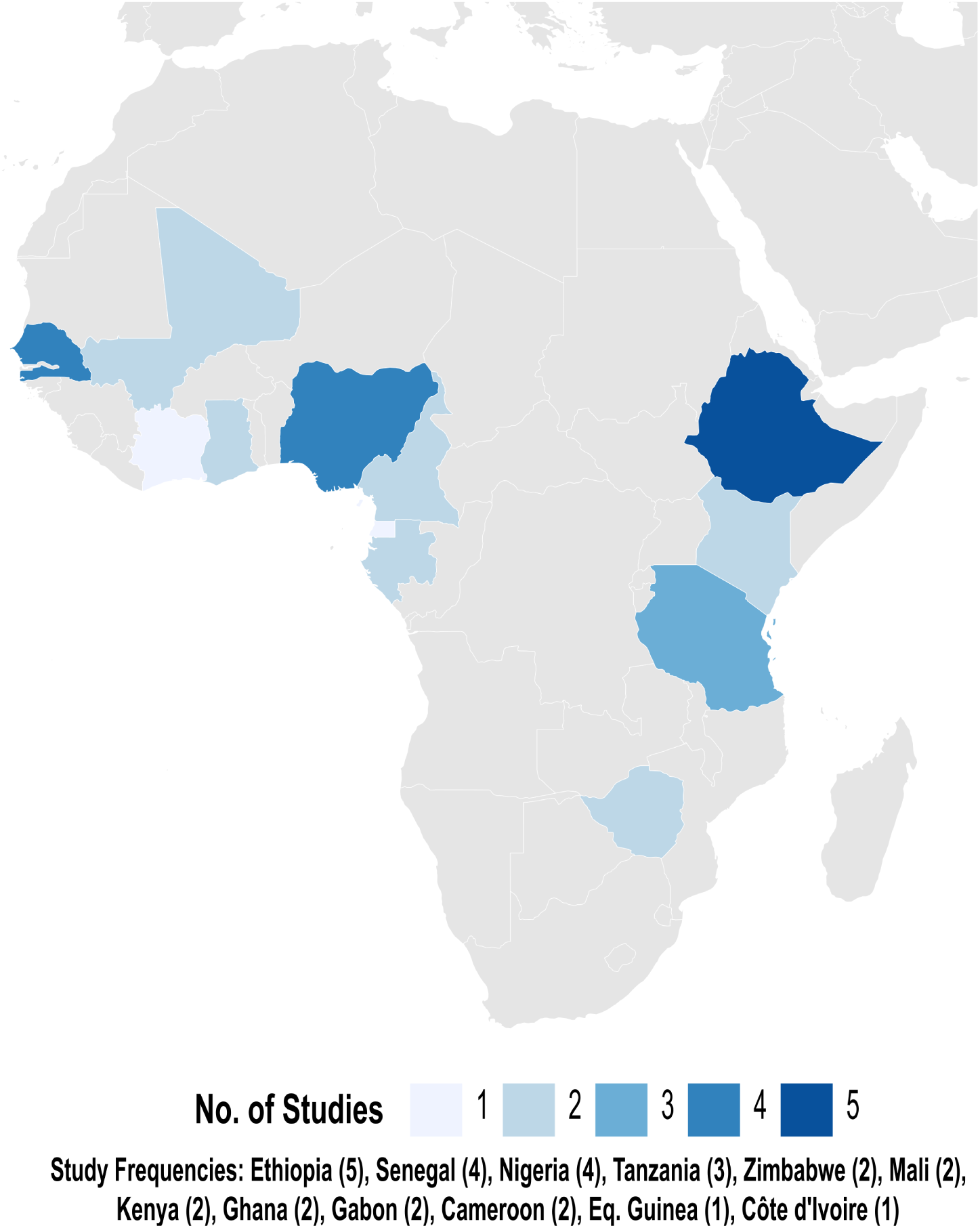
Geographic distribution of included studies. All 30 studies were from 12 countries in Africa. Countries are colored based on the total number of studies.

**Fig 3.**
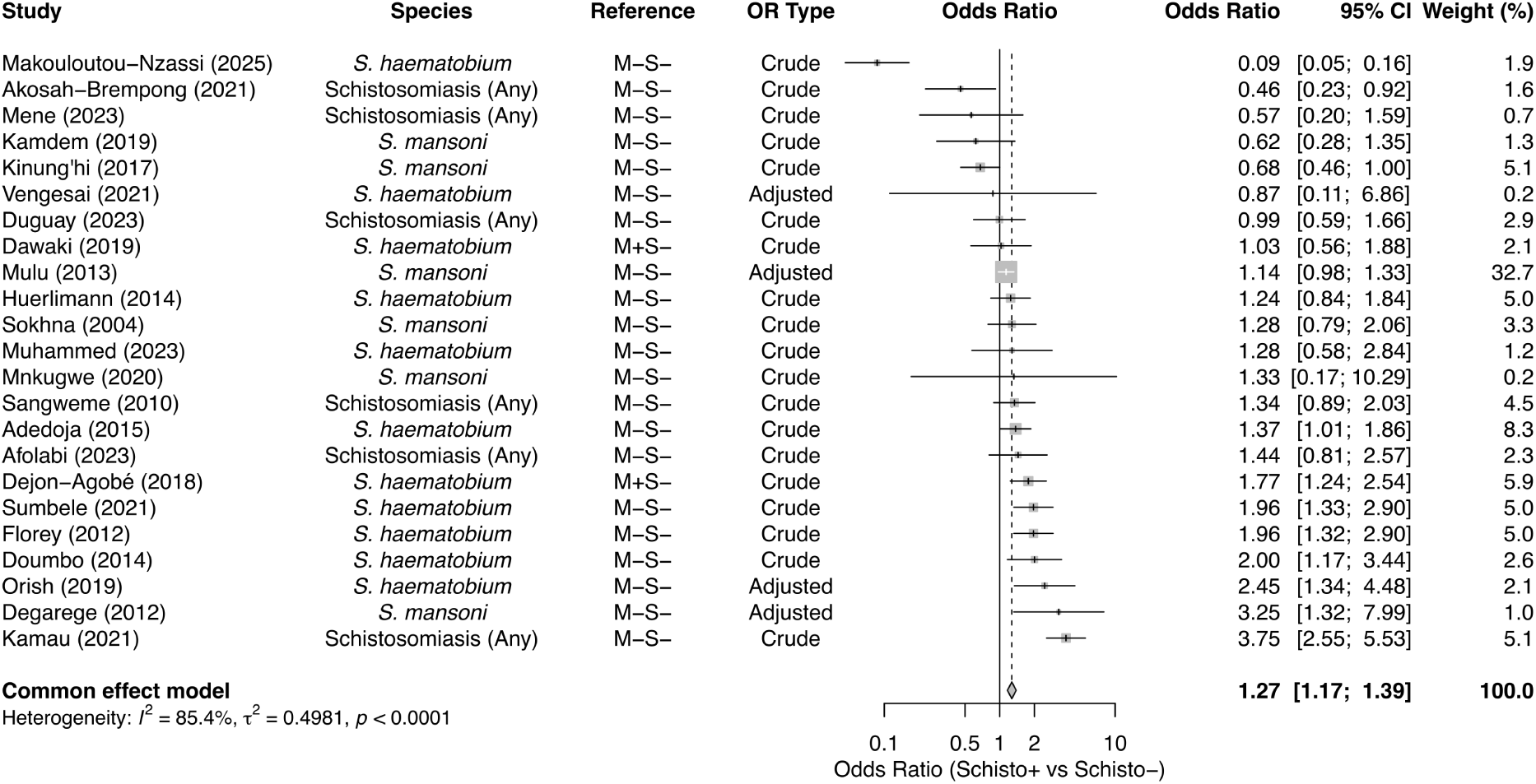
Meta-analysis of the association between schistosomiasis infection and malaria. Overall common-effects model using data from 27 studies. The M-S- and M+S- in the Reference column refer to the reference group used in the primary study (M-S- = uninfected; M+S- = single infection).

When stratified by *Schistosoma* species (Figure 4), the association with malaria remained significant for both species. *S. haematobium* infection was associated with a 46% increase in the odds of malaria (OR 1.46, 95% CI: 1.27–1.67; 13 studies), whereas *S. mansoni* showed a 16% increase (OR 1.16, 95% CI: 1.02–1.31; 11 studies). The difference between species was statistically significant (*p* = 0.0133). The consistency of evidence also varied by species. For *S. haematobium*, eight of the 13 studies (61.5%) reported a statistically significant positive association. In contrast, for *S. mansoni*, three of 11 studies (27.3%, 3/11) reported a significant positive association, while one study reported a significant negative association.

**Fig 4.**
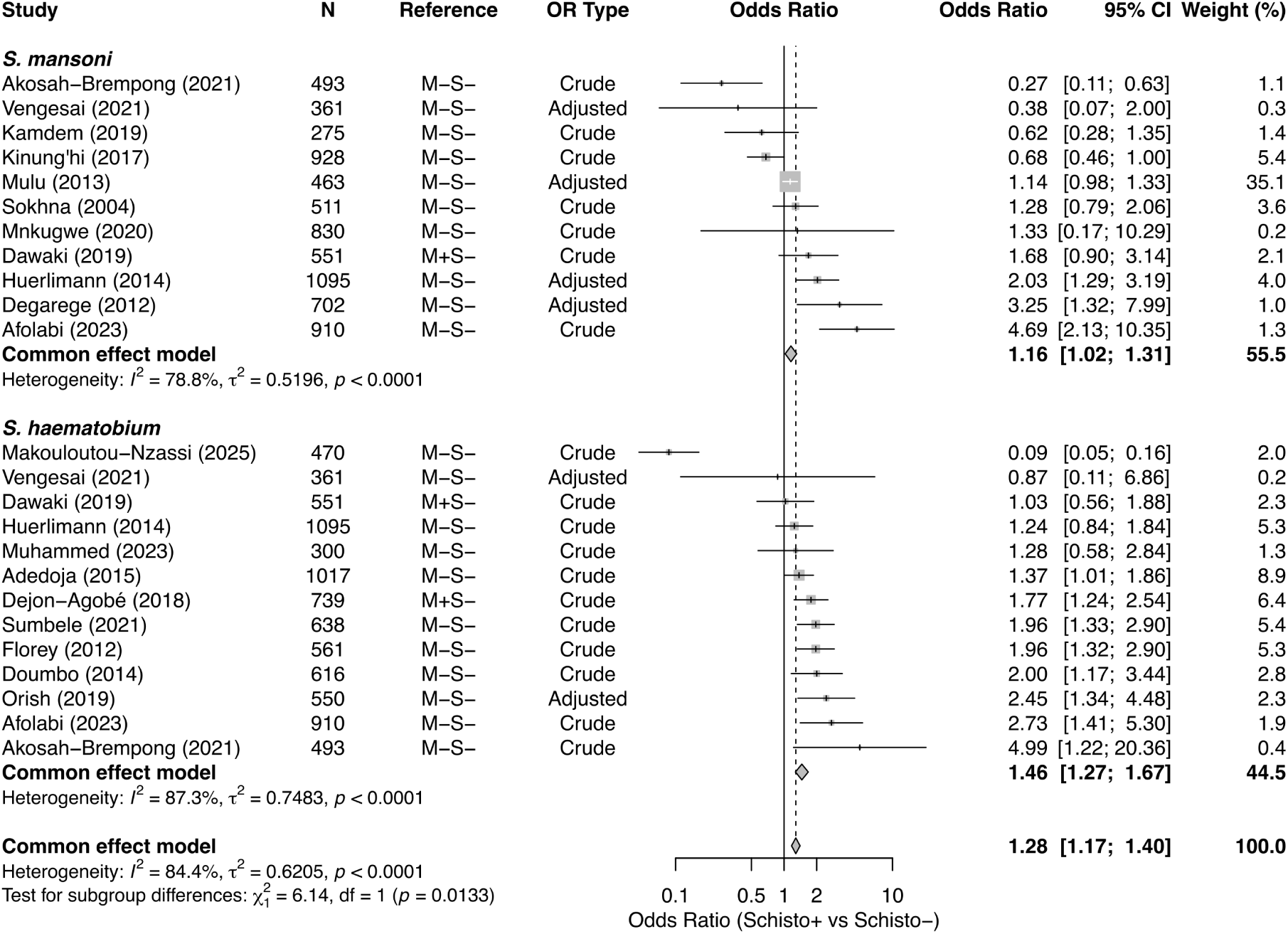
Subgroup meta-analysis by *Schistosoma* species. Common-effects models stratified by *S. mansoni* (9 studies) and *S. haematobium* (11 studies).

When stratified by the reference population (Figure 5), the association remained consistent regardless of the control group definition. The 25 studies using an uninfected population (M-S-) as the reference group showed a pooled OR of 1.25 (95% CI: 1.14–1.37), with high heterogeneity (*I*^2^ = 86.4%). The two studies using a single-infection population (M+S-) as the reference group yielded a slightly higher point estimate (OR 1.53, 95% CI: 1.13–2.09), with moderate heterogeneity (*I*^2^ = 56.7%, *p* = 0.1285). The difference between these subgroups was not statistically significant (*p* = 0.2163).

**Fig 5.**
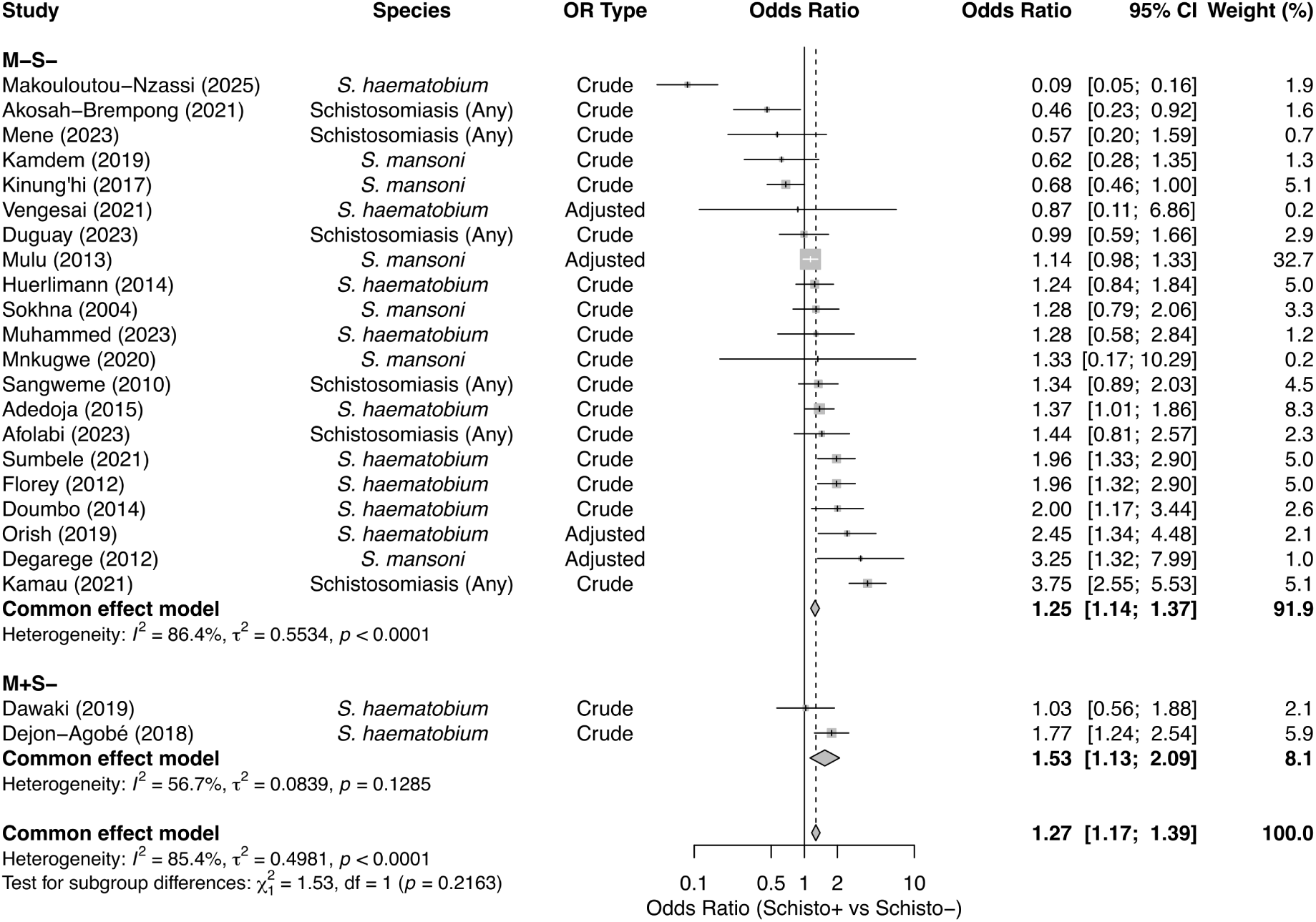
Subgroup meta-analysis by reference population. Common-effects models stratified by primary study reference group: uninfected controls (M-S-; 25 studies) versus single-infection controls (M+S-; 2 studies).

Stratification by study design (Figure 6) showed a statistically significant difference between subgroups (*p* = 0.0408). The association was stronger in the four cohort studies (OR 1.57, 95% CI: 1.26–1.94, *I*^2^ = 0%, *p* = 0.4786). In contrast, while the association remained significant among the 23 cross-sectional studies (OR 1.22, 95% CI: 1.11–1.35), the effect size was smaller and substantial heterogeneity was observed (*I*^2^ = 87.5%, *p <* 0.0001).

**Fig 6.**
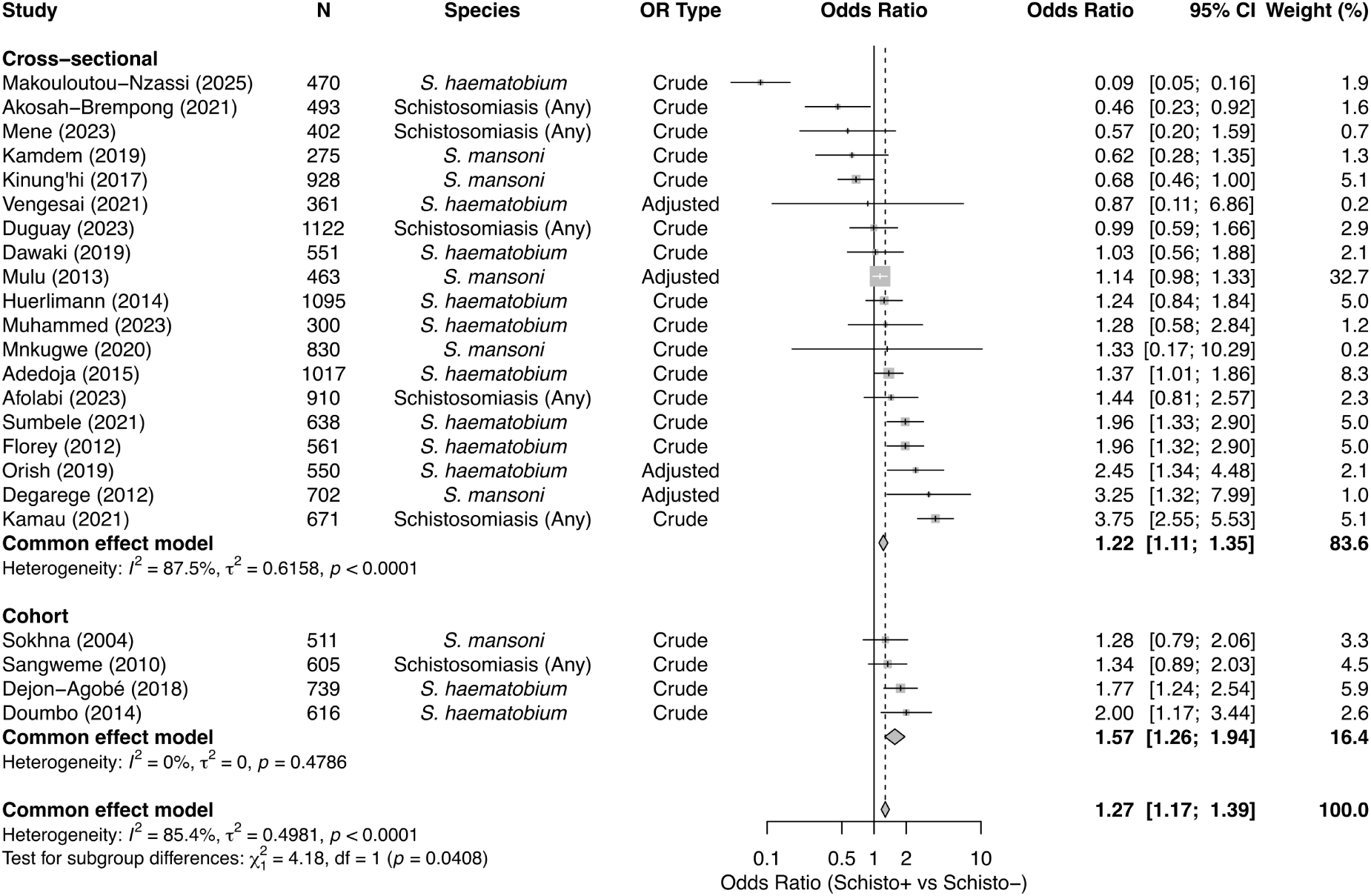
Subgroup meta-analysis by study design. Common-effects models stratified by primary study design: Cross-sectional (23 studies) versus Cohort (4 studies).

Analysis by study setting (Figure 7) revealed significant differences between groups (*p* = 0.0003). The association was strongest in community-based studies (OR 1.68, 95% CI: 1.39–2.04; 6 studies; *I*^2^ = 83.5%) and studies conducted in both communities and schools (OR 1.66, 95% CI: 1.23–2.25; 3 studies), the latter of which showed no heterogeneity (*I*^2^ = 0%). Conversely, no significant association was observed in the 10 school-based studies (OR 1.03, 95% CI: 0.88–1.21), with this subgroup exhibiting substantial heterogeneity (*I*^2^ = 90.3%). Studies conducted in health clinics showed a modest but significant positive association (OR 1.18, 95% CI: 1.02–1.36; 3 studies; *I*^2^ = 61.6%).

**Fig 7.**
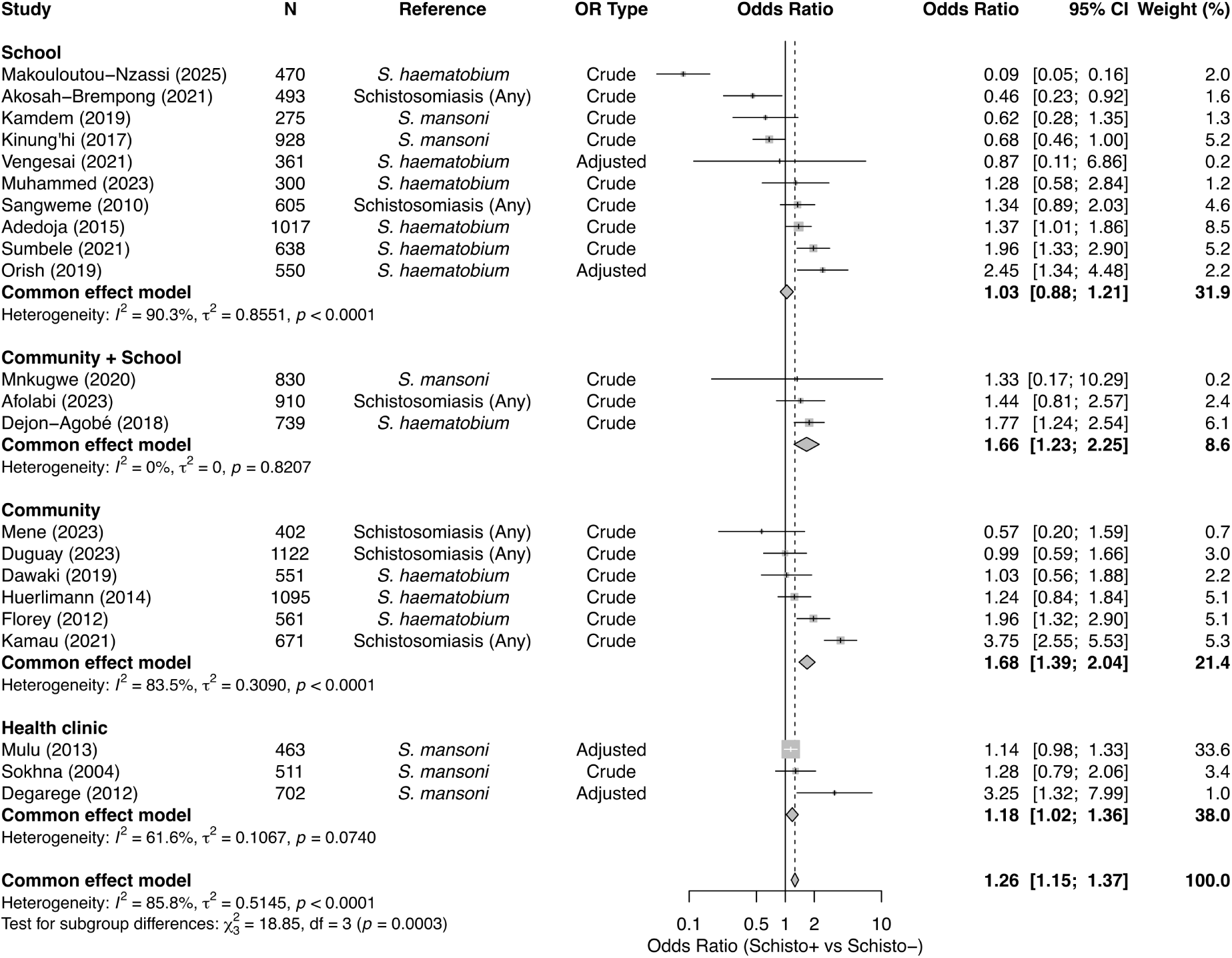
Subgroup meta-analysis by study setting. Common-effects models stratified by setting: School (9 studies), Community + School (3 studies), Community (7 studies), and Health clinic (3 studies).

We also stratified by the type of effect measure provided (Supplementary Figure S3). The positive association remained significant for both studies reporting crude odds ratios (OR 1.30, 95% CI: 1.17–1.45; 22 studies; *I*^2^ = 87.1%) and those reporting adjusted odds ratios (OR 1.22, 95% CI: 1.06–1.41; 5 studies; *I*^2^ = 71.7%). There was no significant difference between adjusted versus unadjusted studies (*p* = 0.4829).

The ROB analysis (Supplementary Figure S4) also showed a significant difference between subgroups (*p* = 0.0099). The positive association was significant for both low-risk studies (OR 1.35, 95% CI: 1.20–1.51; 10 studies; *I*^2^ = 43.4%) and moderate-risk studies (OR 1.16, 95% CI: 1.04–1.29; 16 studies; *I*^2^ = 89.9%). The single high-risk study showed no significant association (OR 1.28, 95% CI: 0.79–2.06).

### Association of sex with co-infection

Thirteen studies were included in the meta-analysis of sex as a risk factor for schistosome-malaria co-infection, with males as the exposure group and females as the reference. The overall pooled analysis showed that males had 1.49 times higher odds of co-infection compared to females (95% CI: 1.26–1.76; Figure 9). High heterogeneity was observed across studies (*I*^2^ = 78.8%, *p <* 0.0001). Assessment of publication bias using a funnel plot (Supplementary Figure S2) and Egger’s regression test showed no evidence of significant asymmetry (*t* = *−*0.41*, df* = 11*, p* = 0.6917).

**Fig 8.**
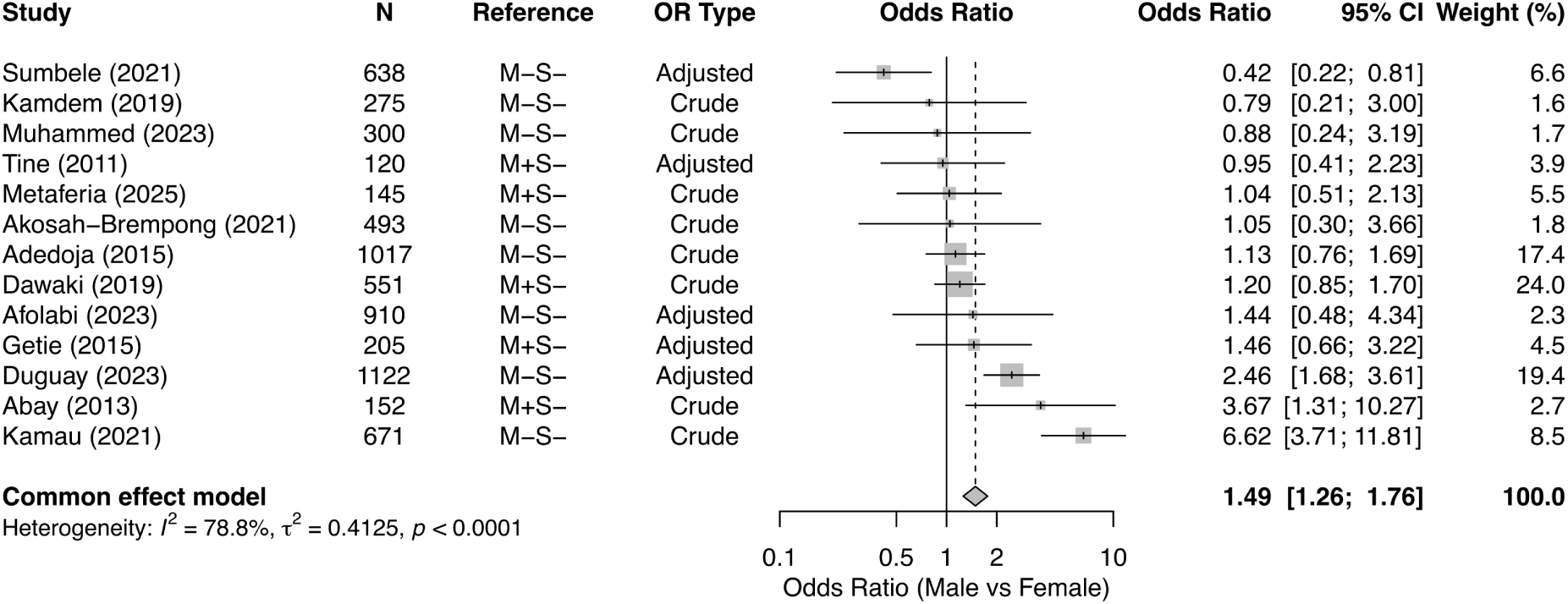
Meta-analysis of the association between sex and co-infection. Overall common-effects model (13 studies) with males as the exposure group.

**Fig 9.**
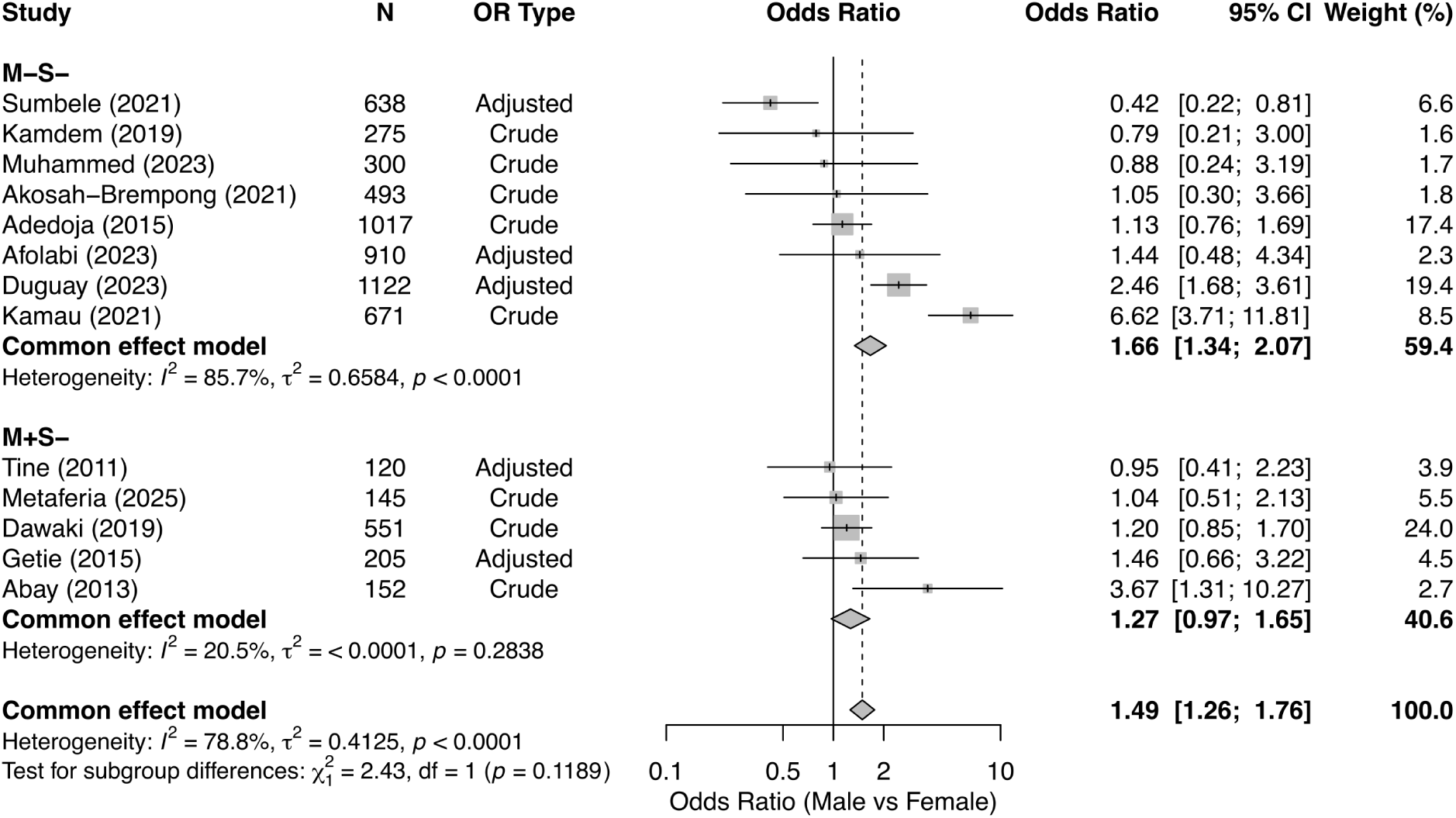
Subgroup meta-analysis of sex by reference population. Common-effects models stratified by primary study reference group: uninfected controls (M-S-; 8 studies) versus single-infection controls (M+S-; 5 studies).

The subgroup analysis of sex based on the reference population for infection is shown in Figure 9. In the eight studies that used an uninfected population (M-S-) as the reference group, males had significantly higher odds of co-infection (OR 1.66, 95% CI: 1.34–2.07). This subgroup showed high heterogeneity (*I*^2^ = 85.7%, *p <* 0.0001). In the five studies using a single-infection population (M+S-) as the reference, the association was not significant (OR 1.27, 95% CI: 0.97–1.65), and heterogeneity was low (*I*^2^ = 20.5%, *p* = 0.2838). There was no statistically significant difference between the two (M-S-, M+S-) subgroups (*p* = 0.1189).

The subgroup analysis for sex by study setting revealed a significant difference between groups (*p <* 0.0001; Figure 10). In three community-based studies, males had 2.08 times higher odds of co-infection than females (95% CI: 1.64–2.63), though with very high heterogeneity (*I*^2^ = 92.3%). Conversely, in five school-based studies, there was no significant association of sex with co-infection (OR 0.87, 95% CI: 0.64–1.19), with moderate heterogeneity (*I*^2^ = 38.1%). The four studies conducted in health clinics also showed no statistically significant association of sex with co-infection (OR 1.37, 95% CI: 0.90–2.07) and had moderate heterogeneity (*I*^2^ = 37.7%).

**Fig 10.**
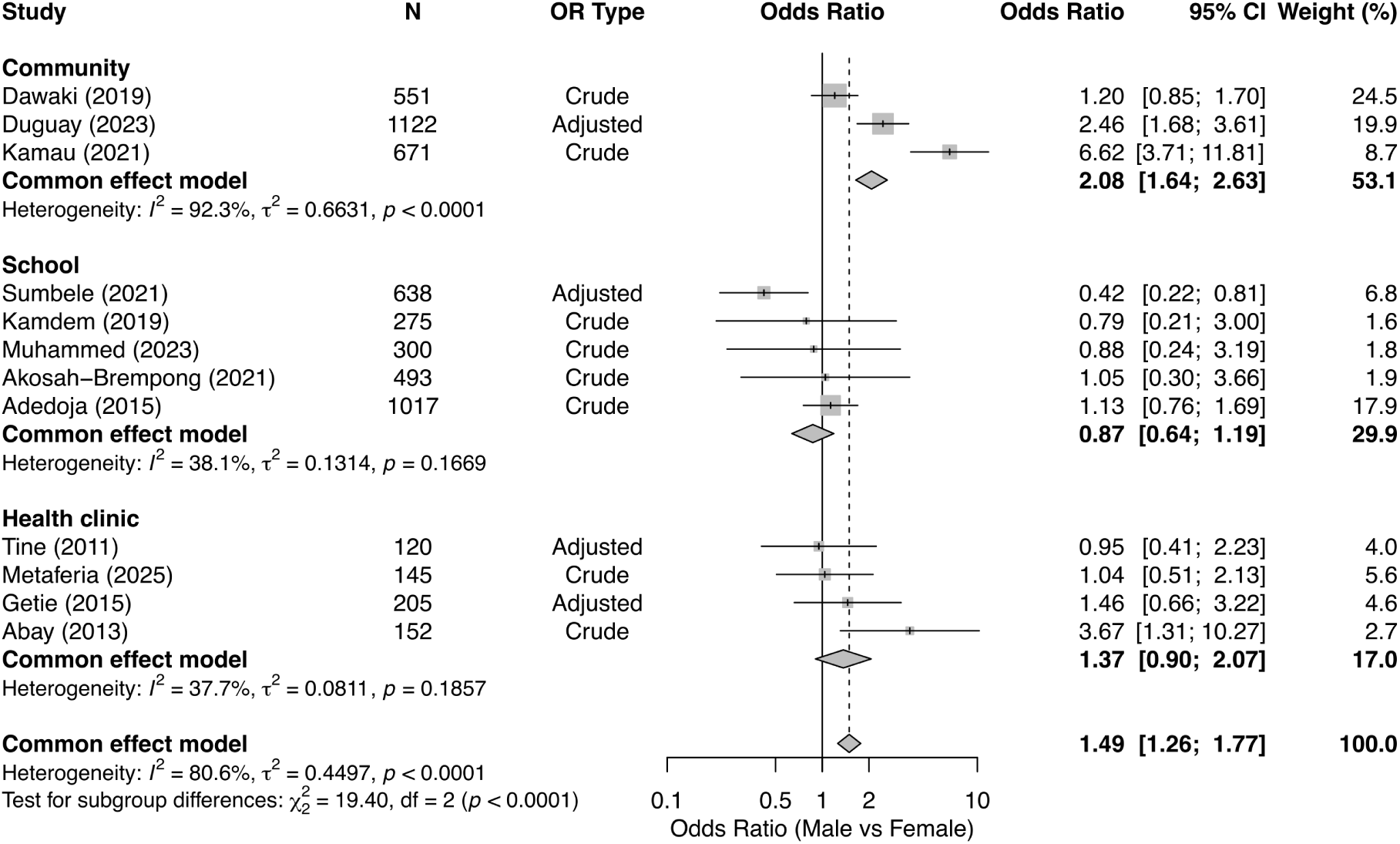
Subgroup meta-analysis of sex by study setting. Common-effects models stratified by setting: Community (3 studies), School (5 studies), and Health clinic (4 studies).

We also stratified the analysis by the type of effect measure provided (see Supplementary Figure S5). The positive association remained significant regardless of study design, showing an adjusted odds ratio of 1.47 (95% CI: 1.10–1.96; four studies; *I*^2^ = 86.4%) and a crude odds ratio of 1.50 (95% CI: 1.22–1.86; eight studies; *I*^2^ = 79.7%). There was no significant difference between these subgroups (*p* = 0.8989). Stratification by the ROB analysis (see Supplementary Figure S6) also showed no significant difference between subgroups (*p* = 0.8548). The positive association was significant for both low-risk studies (OR 1.46, 95% CI: 1.10–1.93; 4 studies; *I*^2^ = 86.3%) and moderate-risk studies (OR 1.51, 95% CI: 1.22–1.87; 8 studies; *I*^2^ = 79.8%).

### Association of water contact with co-infection

Only three studies were available for the meta-analysis of water contact as a risk factor for co-infection. Water contact was associated with 2.53 times higher odds of co-infection than individuals without reported water contact (95% CI: 1.60–4.00; Figure 11). Moderate heterogeneity was observed (*I*^2^ = 55.3%), though it was not statistically significant (*p* = 0.1068).

**Fig 11.**
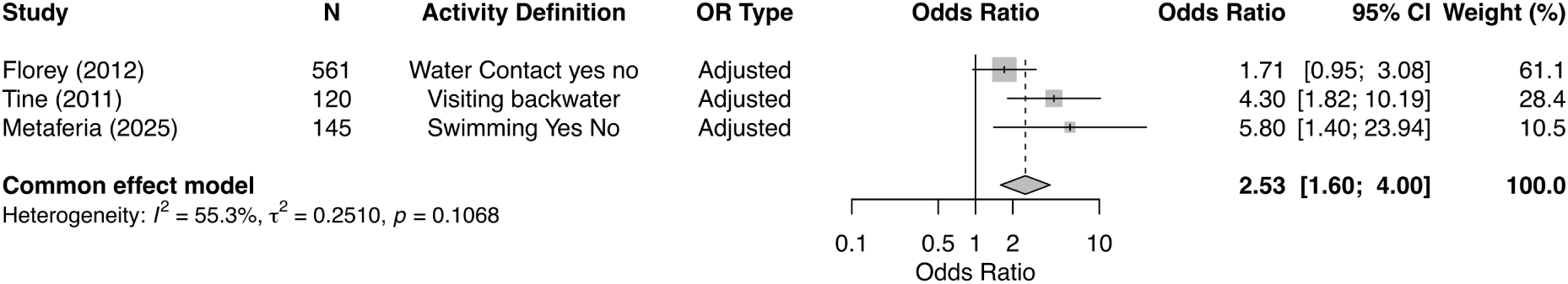
Meta-analysis of the association between water contact and co-infection. Overall common-effects model using data from 3 studies.

### Statistical methodologies and model Setups

The approaches employed to estimate the risk of co-infection varied considerably depending on how the relationship between the parasites was conceptualized. The most common approach (70.0%, 21/30) utilized standard logistic regression. Within this framework, researchers adopted two distinct strategies. The predominant strategy (53.3%, 16/30) treated co-infection as a distinct binary outcome (co-infection vs. no co-infection), effectively modeling the probability of joint carriage as a unique disease state [26, 27]. A second strategy (16.7%, 5/30) modeled single-infection risks, treating *Schistosoma* infection as an independent predictor variable (*X*) for a malaria outcome (*Y* ) often to test specific hypotheses regarding susceptibility [28]. Longitudinal designs employed time-to-event analyses. Cox proportional hazards models were used in two cohort studies to evaluate baseline co-infection status as a predictor for the incidence of febrile malaria episodes [29, 30]. Additionally, continuous clinical outcomes, such as parasite density or hemoglobin levels, were analyzed using linear mixed models [31] or multivariable Poisson regression [5].

Despite the focal nature of both diseases, accounting for spatial clustering was rare. Only three studies (10.0%, 3/30) explicitly accounted for the hierarchical structure of the data (e.g., individuals nested within households or villages) using Mixed-Effect Logistic Regression or Generalized Estimating Equations (GEE) [12, 9, 5]. The remaining 27 studies (90.0%) relied on standard models that assume independence between observations, potentially underestimating standard errors in clustered study designs.

The adjustment for confounding factors was highly variable across the literature. While sociodemographic factors were nearly universal with age included in 93.3% (28/30) and sex in 90.0% (27/30) of models, environmental and behavioral adjustments were inconsistent. Furthermore, despite eight studies collecting data on Long-Lasting Insecticidal Nets (LLIN) ownership, only two formally included it as a covariate in the final model [12, 9]. The method of variable selection was predominantly data-driven. The majority of studies reported using a stepwise approach or univariate screening process, where candidate variables were only retained for the final model if they achieved a statistical threshold (typically *p <* 0.20 or *p <* 0.25) in bivariate analysis [12, 32].

### Narrative synthesis of age, socioeconomic status, and prevention tools

Age was a consistent predictor of co-infection, however reporting categories differed. The majority of studies indicate a peak burden within school-aged children (approximately 6–14 years). For instance, Abay et al. [8] and Duguay et al. [12] identified peak risks in the 6–14 and 5–14 year groups, respectively, with Duguay et al. noting a 20% increase in odds for every one-year increase in age. Florey et al. [33] utilized a broader category (8–17 years) but similarly identified elevated odds in this younger demographic compared to adults. However, this age distribution was not consistent, as Getie et al. [11] reported an older peak, with the highest prevalence (32.5%) observed in young adults aged 16–20 years.

Socioeconomic status and education exhibited a negative association with co-infection. Household infrastructure acted as a proxy for socioeconomic vulnerability; living in houses with mud floors or thatched roofs was associated with higher co-infection rates compared to those with cement floors or corrugated iron roofs [32, 33]. Abay et al. [8] noted a gradient where co-infection rates declined as maternal educational level increased. Similarly, Duguay et al. [12] found that specific knowledge regarding schistosomiasis transmission was protective (aOR 0.71). WASH practices were important, as access to improved water sources and the avoidance of open defecation were negatively associated with co-infection [34].

Evidence regarding malaria prevention tools was partially captured through different definitions. Duguay et al. [12] found that inadequate access to LLINs increased co-infection risk (aOR 1.67) and Florey et al. [33] observed that reported bednet use was associated with lower odds of co-infection.

## Discussion

Integrating control programs for malaria and neglected tropical diseases, such as schistosomiasis, is a key global health strategy advocated by the World Health Organization (WHO) [35]. This systematic review and meta-analysis synthesized evidence from 30 studies across sub-Saharan Africa involving 16,775 participants. Our findings reveal that the risk profile for co-infection is multi-dimensional, driven by an interplay of sociodemographic and environmental factors. While sociodemographic dimensions, particularly age and sex, were the most commonly assessed, we found that the evidence for other dimensions, such as specific behaviors, environmental exposures, and socioeconomic status, is less developed, highlighting a major gap in the current understanding of co-infection risk.

Our meta-analysis shows a significant positive association between schistosomiasis and malaria, a finding that remained robust in both crude and adjusted analyses. This association was significant for both *S. haematobium* (OR 1.46) and *S. mansoni* (OR 1.16). It is critical, however, to interpret this primarily as a measure of co-occurrence rather than a directional risk. Visual inspection of the forest plots suggests that this overall effect is heavily driven by a subset of large studies in *S. haematobium* endemic areas. A plausible explanation for this strong co-occurrence is shared ecological and behavioral confounding rather than immunological synergy. Schistosome transmission is inextricably linked to water contact activities like swimming or bathing [36, 26, 23]. These same stagnant or slow-moving water bodies (e.g., dams, ponds) are ideal breeding sites for *Anopheles* mosquitoes [37, 38]. Thus, individuals performing these activities are simultaneously exposed to both cercariae and infectious mosquito bites, which may generate the observed association.

While our meta-analysis showed a larger effect size for *S. haematobium* than for *S. mansoni*, this apparent species-specific difference should be interpreted with caution. This discrepancy is likely an artifact of the available evidence base. A substantial portion of the included studies were conducted in areas predominantly or exclusively endemic for *S. haematobium* (e.g., in parts of Nigeria and Senegal) and thus did not even assess *S. mansoni* [39, 36, 40]. The geographic and methodological imbalance, combined with variations in diagnostic methods, likely inflates the pooled estimate for *S. haematobium*. Furthermore, biological hypotheses for such a strong difference are contested, as both species are known to induce a potent, systemic Th2-biased immune response [41, 42, 43, 44] and both contribute to anemia, which compounds the hemolytic burden of malaria [36].

Male sex was identified as a significant predictor of co-infection, yet this association is likely driven less by biological susceptibility than by patterns of environmental exposure that emerge during the transition to adulthood. Our subgroup analysis revealed a divergence by study setting. While sex was not a significant risk factor in school-based studies, it was a strong predictor in community-based settings with over 2.08 times higher odds. This aligns with the peak risk window we identified in older children and adolescents, where behavioral patterns begin to diverge along gender lines. In the wider community, the direction of risk appears highly context-specific, reflecting local divisions of labor. In settings where herding and occupational fishing are male-dominated, males bear the burden of co-infection; Duguay et al. [12], Kamau et al. [5], and Abay et al. [8] all reported significantly higher rates in males. These activities frequently require men to remain outdoors near water bodies exposed to *Anopheles* mosquitoes, therefore increasing exposure to both vectors. Conversely, in settings where domestic water contact drives exposure, the risk reverses. Sumbele et al. [34] in Cameroon identified being female as the sole significant risk factor (aOR 2.38), explicitly attributing this to the gendered responsibility for laundry and dishwashing in infested streams. Thus, the observed sex associations likely represent a shared risk profile where specific, context-dependent behaviors increase exposure to both vectors simultaneously.

Our meta-analysis identified water contact as a strong risk factor for co-infection where individuals with water contact had over 2.53 times higher odds than individuals without water contact, potentially representing a shared exposure pathway. However, this quantitative synthesis was restricted to the three studies that provided extractable effect estimates. The broader interpretation of this risk is complicated by the scarcity of data and heterogeneity in measurement. Only five of 30 studies explicitly assessed direct water contact (e.g., swimming, fishing), while a further six studies relied on environmental proxies such as proximity to water bodies. This lack of standardization mirrors findings from a recent review by Reitzug et al. [23], who previously noted inconsistent definitions of water contact. Consequently, with only five studies explicitly adjusting for direct water contact, many estimates in the literature likely suffer from unmeasured confounding. Furthermore, the standard univariate modeling techniques employed in the majority of studies cannot dissect the nature of shared risk. Because water contact is a necessary requirement for schistosome transmission but only a proxy indicator for malaria transmission (via vector proximity). Univariate models that lump co-infection as a binary outcome cannot distinguish if water contact is a true driver of dual susceptibility or if the association is a statistical artifact driven by the effect of water contact on schistosomiasis alone.

Other studies in our review relied on environmental proxies for exposure, finding that living near a river or dam was a consistent risk factor [32], whereas increasing distance from water bodies conferred protection [33]. However, we were unable to synthesize these findings quantitatively due to heterogeneity in measurement scales. For instance, some studies utilized continuous distance metrics, while others relied on site-specific categorical variables, making a pooled effect estimate impossible. To enable future synthesis, the field could adopt standardized spatial methods that are well-established in mono-infection research. The increasing availability of open-access, high-resolution satellite data offers an alternative [45, 46], as researchers may calculate environmental indices such as the Normalized Difference Vegetation Index (NDVI) and Normalized Difference Water Index (NDWI) at granular scales.

Beyond characterizing these environmental drivers of transmission, three studies established the interaction between pathogens as a result of co-infection. Although excluded from our quantitative synthesis due to their focus on continuous clinical outcomes, three studies provide evidence regarding the interaction between *S. haematobium* and *P. falciparum*. Briand et al. [31] reported that children with light *S. haematobium* infections had lower *P. falciparum* densities, while Lyke et al. [43] observed that co-infected children aged 4–8 years experienced fewer clinical malaria episodes and a delayed time to first infection. Conversely, Oboh-Imafidon et al. [40] focused on the modulation of disease severity, suggesting that host genetic factors (such as CD14 variants) play a critical role in determining the intensity of infection (egg counts) in co-infected individuals.

### Limitations

Our review is subject to limitations arising from the review process, measurement inconsistencies, and the methodology of the primary literature. Our search strategy was restricted to studies with titles and abstracts in English. This may have inadvertently excluded relevant local literature, particularly from Francophone regions in sub-Saharan Africa where schistosomiasis is highly endemic. We also observed a high heterogeneity across most meta-analyses (*I*^2^ *>* 80%). While we explored this through subgroup analyses by species, setting, and study design, considerable residual heterogeneity remained. Limitations regarding exposure definitions in the primary studies also constrain our findings. For water contact specifically, definitions varied, ranging from broad environmental proxies (e.g., ”living near water”) to specific behaviors (e.g., ”swimming”). Further, this review is limited by the reliance on standard logistic regression to model co-infection as a simple binary outcome (co-infected vs. not co-infected). By collapsing the four distinct infection states (uninfected, malaria-only, schistosomiasis-only, and co-infected) into a dichotomy, these models fail to simultaneously estimate the probabilities of each parasite and the correlation between them [17]. This leads to ambiguity regarding the reference population; comparing ”co-infected” individuals to a ”not co-infected” group compares uninfected individuals with those harboring single infections, potentially biasing risk estimates if the single-infection groups have divergent risk profiles. Finally, the predominant cross-sectional nature of the evidence base precludes inference regarding the sequence of parasite acquisition. Without longitudinal data, it remains impossible to determine if prior schistosomiasis infection alters susceptibility to subsequent malaria through immunomodulation, or to answer dynamic clinical questions, such as whether the order of infection influences the severity of morbidity.

## Conclusion

This systematic review shows that schistosomiasis and malaria share a complex risk profile driven by overlapping sociodemographic and endogenous factors. While age and sex are well-established determinants, our synthesis reveals a scarcity of standardized evidence regarding behavioral, environmental, and socioeconomic dimensions. Current evidence is heavily reliant on cross-sectional studies and univariate models, which cannot capture the dynamic nature of co-infection. To move from separate vertical programs to the integrated interventions envisioned in the WHO Neglected Tropical Disease Roadmap 2030 [35], future research must prioritize longitudinal designs and standardized, scalable environmental metrics. Our review suggests that targeted interventions should be focused on older children and adolescents, and possibly males in high-risk community settings.

## List of Abbreviations

Abbreviation: Definition
aOR: Adjusted Odds Ratio
CI: Confidence Interval
cOR: Crude Odds Ratio
GEE: Generalized Estimating Equations
HR: Hazard Ratio
LLIN: Long-Lasting Insecticidal Net
MeSH: Medical Subject Heading
NDVI: Normalized Difference Vegetation Index
NDWI: Normalized Difference Water Index
NIH: National Institutes of Health
OR: Odds Ratio
PCR: Polymerase Chain Reaction
PCV: Packed Cell Volume
POC-CCA: Point-of-Care Circulating Cathodic Antigen
PRISMA: Preferred Reporting Items for Systematic Reviews and Meta-Analyses RDT Rapid Diagnostic Test
ROB: Risk of Bias
RR: Relative Risk
sRDT: Schistosome Rapid Diagnostic Test
WASH: Water, Sanitation, and Hygiene
WHO: World Health Organization

## Declarations

## Data Availability

All data in the present work are contained in the manuscript and supplementary files.

## Acknowledgments

We thank Nia Roberts and Dr Reem Malouf for supporting the initial review stages, and acknowledge Fabian Reitzug and Zhaoyu Guo for providing translations of non-English articles. We also thank the Chami Group for their feedback during group meetings.

## Author contribution

Conceptualization: MML, GFC Data curation: MML and BL Analysis: MML

Funding acquisition: MML and GFC Methodology: MML

Resources: GFC Supervision: GFC and CAD

Writing – original draft: MML

Writing – review and editing: MML, BL, CAD, and GFC

