## Supplementary material for "Shared risk factors for malaria and schistosomiasis co-infection: a systematic review and meta-analysis"

### Supporting information

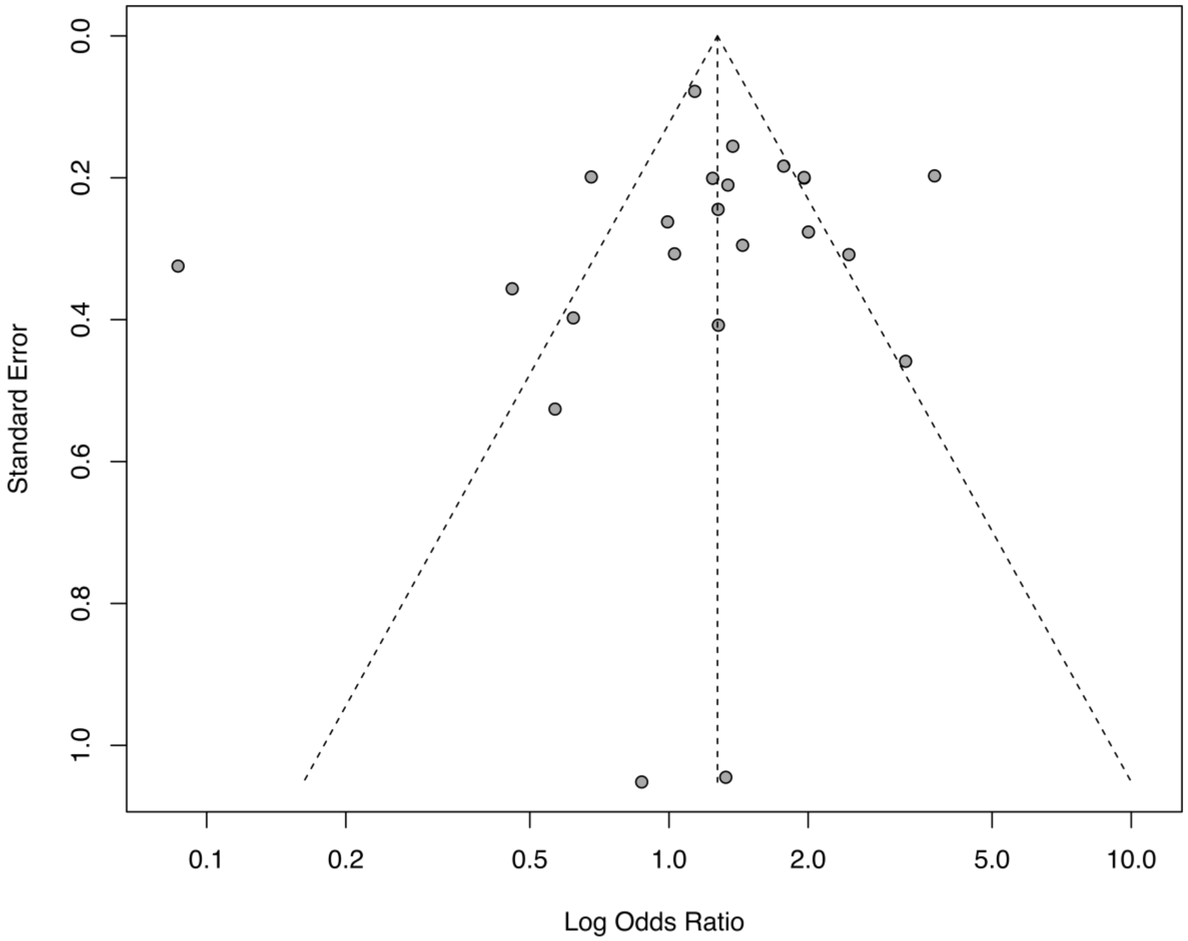


**Fig S1. Funnel plot for schistosomiasis as a risk factor.** Plot of log odds ratio versus standard error for studies included in the meta-analysis for the association between schistosomiasis and malaria.

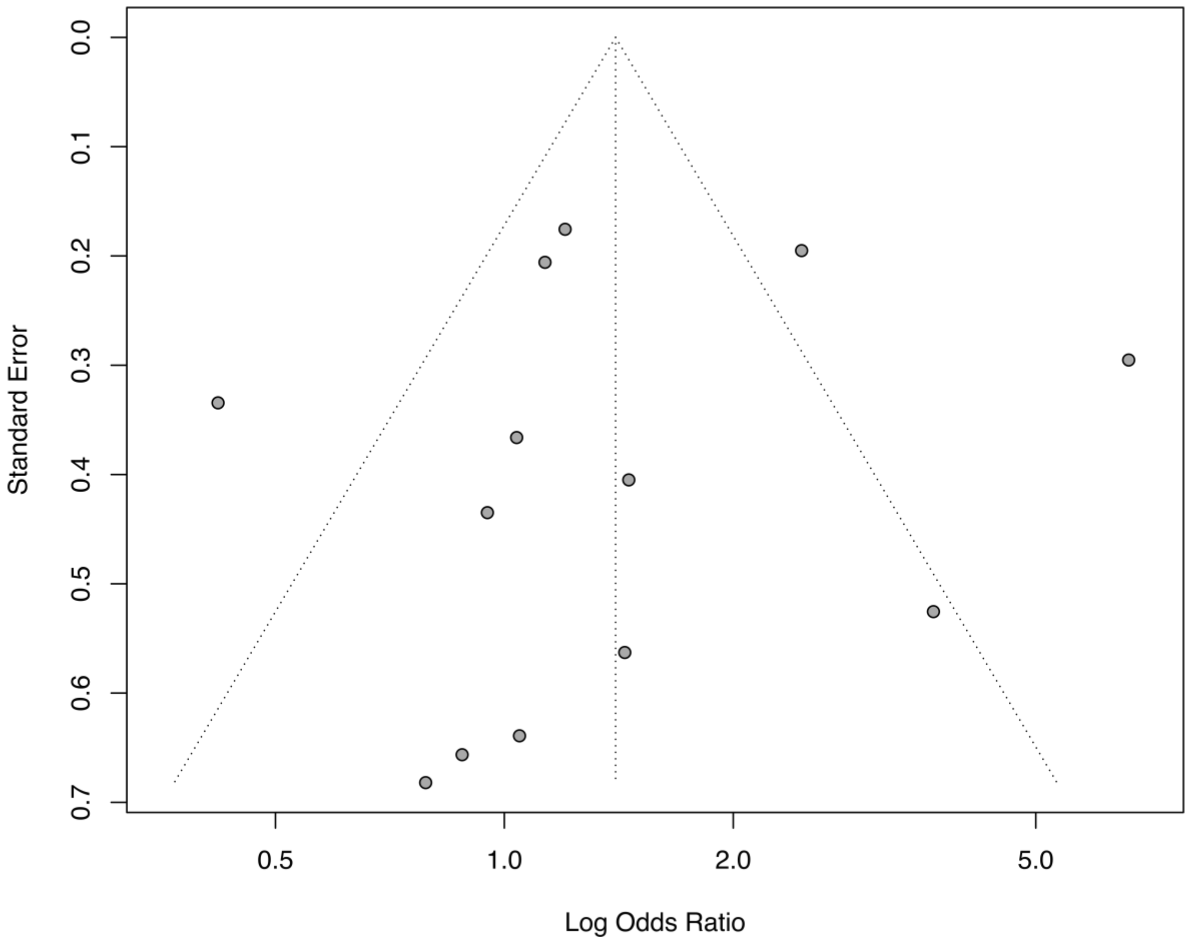


**Fig S2.** Funnel plot for sex as a risk factor. Plot of log odds ratio versus standard error for studies included in the meta-analysis for the association between sex and co-infection.

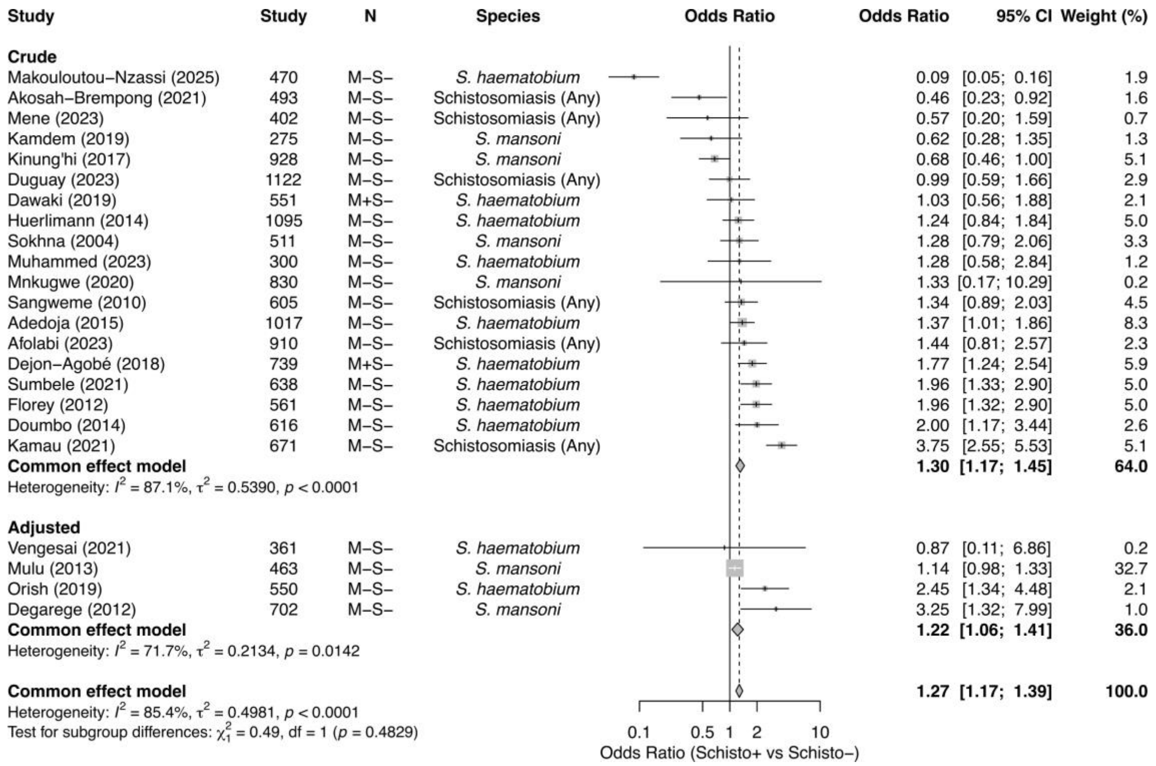


**Fig S3.** Subgroup meta-analysis by effect estimate type. Common-effects models stratified by crude versus adjusted odds ratios.

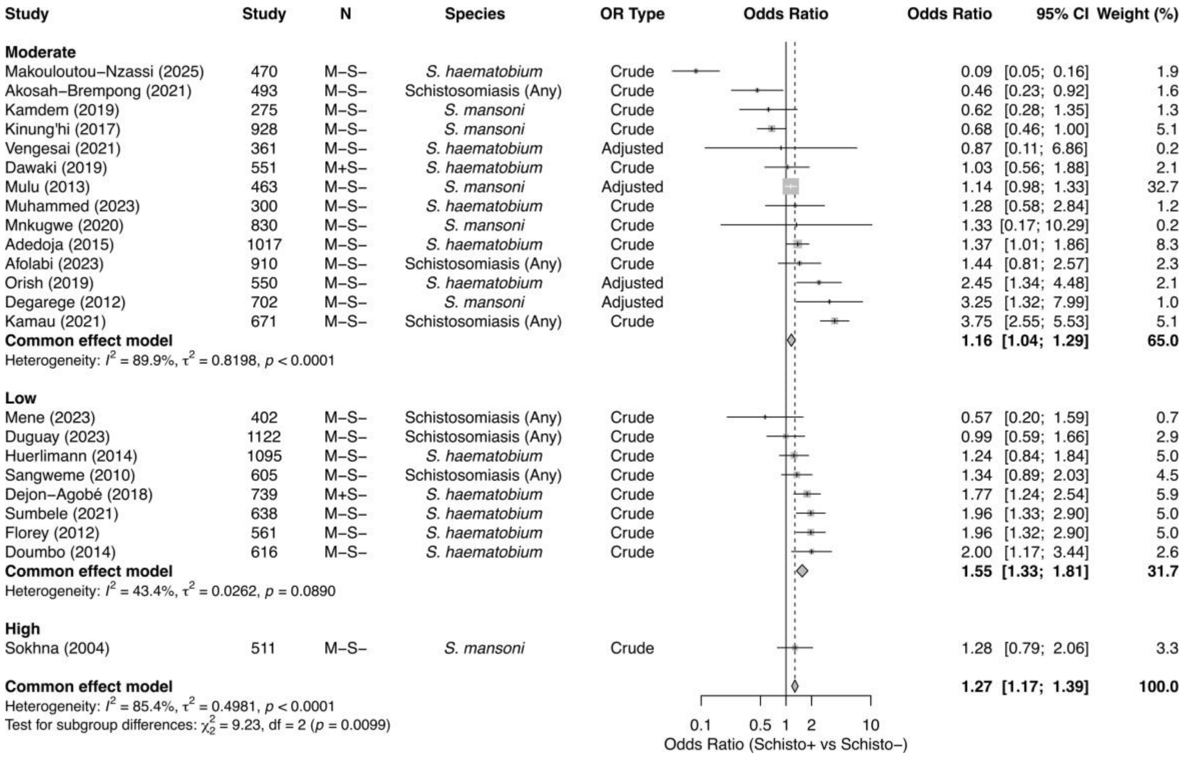


**Fig S4.** Subgroup meta-analysis by ROB. Common-effects models stratified by ROB rating (Low, Moderate, High).

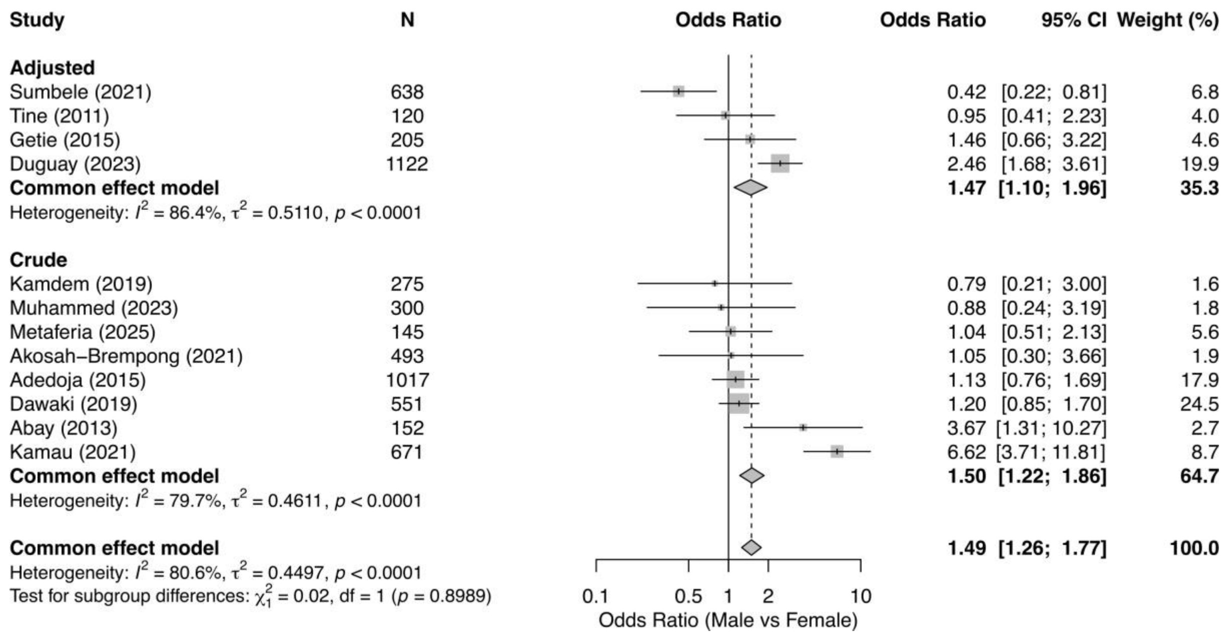


**Fig S5. Subgroup meta-analysis of sex by effect estimate type.** Common-effects models stratified by adjusted (4 studies) versus crude (8 studies) odds ratios.

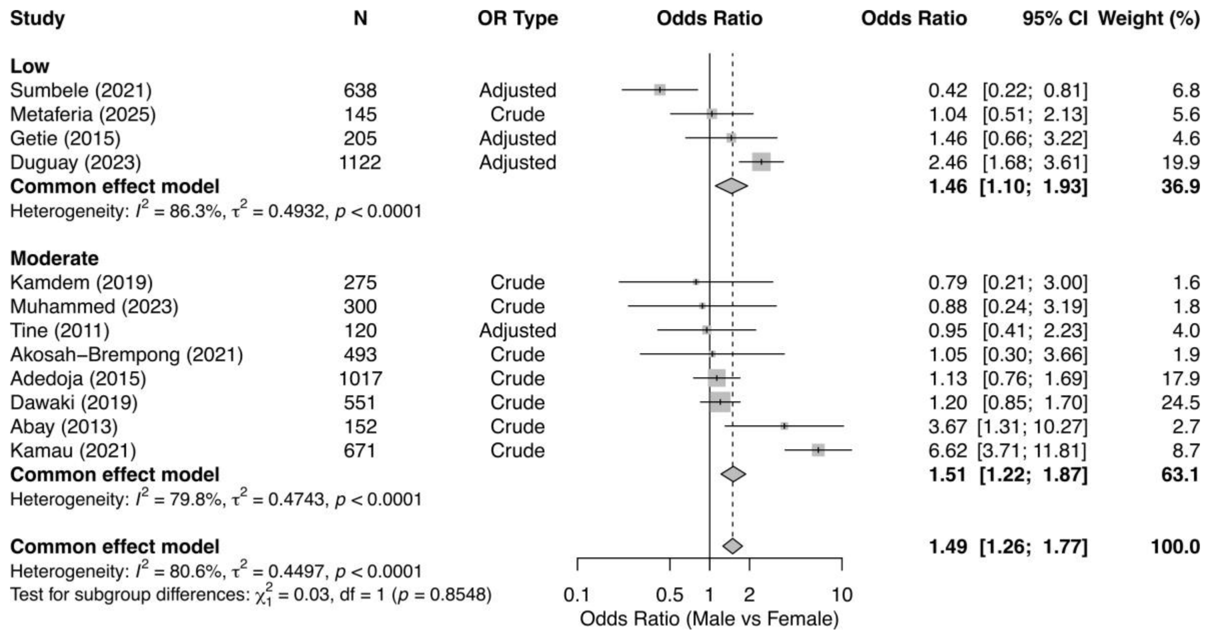


**Fig S6. Subgroup meta-analysis of sex by ROB.** Common-effects models stratified by ROB rating: Low (4 studies) and Moderate (8 studies).

S1 Dataset. Data Extraction Table including study characteristics and full references

S2 Dataset. Risk of Bias Assessment results

S1 Text. Search Strategy

Logic of the search string is as follows (categories connected by ‘AND’):

| **Disease 1** | **Disease 2** | **Co-Infection** |
|---|---|---|
| (malaria OR Plasmodium OR Plasmodium falciparum OR “P. falciparum” OR “Plasmodium vivax" OR malarial parasite) | (schistosom* OR schistosomiasis OR schistosoma OR schistosome OR bilharzia* OR Schistosoma mansoni OR Schistosoma haematobi* OR helminthiasis OR snail fever) | (co-infection* OR "dual infection" OR "co-occurrence" OR cooccur* OR “co-distributed” OR “co-distribution” OR codistribut* OR "concomitant" OR polyparasitism OR multiparasitism OR multi-parasitism OR multi-infection OR multiinfection OR “simultaneous infection” OR “sequential infection “OR joint infection OR superinfection OR synergistic infection) |

The generic search string was adapted to suit constraints and functionalities of different databases. Medline, Embase and Global Health were searched through Ovid (https://www.wolterskluwer.com/en/solutions/ovid). Web of Science was accessed through Clarivate (https://clarivate.com).

OVID (703 Results)

Medline 192

| # | Query | Results from 19 Feb 2025 |
|---|---|---|
| ***1*** | (malaria or Plasmodium or Plasmodium falciparum or "P. falciparum" or "Plasmodium vivax" or malarial parasite).mp. [mp=title, book title, abstract, original title, name of substance word, subject heading word, floating sub-heading word, keyword heading word, organism supplementary concept word, protocol supplementary concept word, rare disease supplementary concept word, unique identifier, synonyms, population supplementary concept word, anatomy supplementary concept word] | 125,708 |
| ***2*** | (schistosom* or schistosomiasis or schistosoma or schistosome or bilharzia* or Schistosoma mansoni or Schistosoma haematobi* or helminthiasis or snail fever).mp. [mp=title, book title, abstract, original title, name of substance word, subject heading word, floating sub-heading word, keyword heading word, organism supplementary concept word, protocol supplementary concept word, rare disease supplementary concept | 48,617 |

| | | |
|---|---|---|
| | word, unique identifier, synonyms, population supplementary concept word, anatomy supplementary concept word] | |
| 3 | (co-infection* or "dual infection" or "co-occurrence" or cooccur* or "co-distributed" or "co-distribution" or codistribut* or "concomitant" or polyparasitism or multiparasitism or multi-parasitism or multi-infection or multiinfection or "simultaneous infection" or "sequential infection OR joint infection" or superinfection or synergistic infection).mp. [mp=title, book title, abstract, original title, name of substance word, subject heading word, floating sub-heading word, keyword heading word, organism supplementary concept word, protocol supplementary concept word, rare disease supplementary concept word, unique identifier, synonyms, population supplementary concept word, anatomy supplementary concept word] | 275,213 |
| 4 | 1 and 2 and 3 | 192 |

Embase 345

| # | Query | Results from 19 Feb 2025 |
|---|---|---|
| ***1*** | (malaria or Plasmodium or Plasmodium falciparum or "P. falciparum" or "Plasmodium vivax" or malarial parasite).mp. [mp=title, book title, abstract, original title, name of substance word, subject heading word, floating sub-heading word, keyword heading word, organism supplementary concept word, protocol supplementary concept word, rare disease supplementary concept word, unique identifier, synonyms, population supplementary concept word, anatomy supplementary concept word] | 156408 |
| ***2*** | (schistosom* or schistosomiasis or schistosoma or schistosome or bilharzia* or Schistosoma mansoni or Schistosoma haematobi* or helminthiasis or snail fever).mp. [mp=title, book title, abstract, original title, name of substance word, subject heading word, floating sub-heading word, keyword heading word, organism supplementary concept word, protocol supplementary concept word, rare disease supplementary concept word, unique identifier, synonyms, population supplementary concept word, anatomy | 53670 |

| | | |
|---|---|---|
| | supplementary concept word] | |
| 3 | (co-infection* or "dual infection" or "co-occurrence" or cooccur* or "co-distributed" or "co-distribution" or codistribut* or "concomitant" or polyparasitism or multiparasitism or multi-parasitism or multi-infection or multiinfection or "simultaneous infection" or "sequential infection OR joint infection" or superinfection or synergistic infection).mp. [mp=title, book title, abstract, original title, name of substance word, subject heading word, floating sub-heading word, keyword heading word, organism supplementary concept word, protocol supplementary concept word, rare disease supplementary concept word, unique identifier, synonyms, population supplementary concept word, anatomy supplementary concept word] | 396025 |
| 4 | 1 and 2 and 3 | 345 |

Global Health 166

| # | Query | Results from 19 Feb 2025 |
|---|---|---|
| ***1*** | (malaria or Plasmodium or Plasmodium falciparum or "P. falciparum" or "Plasmodium vivax" or malarial parasite).mp. [mp=title, book title, abstract, original title, name of substance word, subject heading word, floating sub-heading word, keyword heading word, organism supplementary concept word, protocol supplementary concept word, rare disease supplementary concept word, unique identifier, synonyms, population supplementary concept word, anatomy supplementary concept word] | 106582 |
| ***2*** | (schistosom* or schistosomiasis or schistosoma or schistosome or bilharzia* or Schistosoma mansoni or Schistosoma haematobi* or helminthiasis or snail fever).mp. [mp=title, book title, abstract, original title, name of substance word, subject heading word, floating sub-heading word, keyword heading word, organism supplementary concept word, protocol supplementary concept word, rare disease supplementary concept word, unique identifier, synonyms, population supplementary concept word, anatomy | 39488 |

| | | |
|---|---|---|
| | supplementary concept word] | |
| 3 | (co-infection* or "dual infection" or "co-occurrence" or cooccur* or "co-distributed" or "co-distribution" or codistribut* or "concomitant" or polyparasitism or multiparasitism or multi-parasitism or multi-infection or multiinfection or "simultaneous infection" or "sequential infection OR joint infection" or superinfection or synergistic infection).mp. [mp=title, book title, abstract, original title, name of substance word, subject heading word, floating sub-heading word, keyword heading word, organism supplementary concept word, protocol supplementary concept word, rare disease supplementary concept word, unique identifier, synonyms, population supplementary concept word, anatomy supplementary concept word] | 43353 |
| 4 | 1 and 2 and 3 | 166 |

(malaria OR Plasmodium OR Plasmodium falciparum OR “P. falciparum” OR “Plasmodium vivax" OR malarial parasite)
AND
(schistosom* OR schistosomiasis OR schistosoma OR schistosome OR bilharzia* OR Schistosoma mansoni OR Schistosoma haematobi* OR helminthiasis OR snail fever)
AND
(co-infection* OR "dual infection" OR "co-occurrence" OR cooccur* OR “co-distributed” OR “co-distribution” OR codistribut* OR "concomitant" OR polyparasitism

OR multiparasitism OR multi-parasitism OR multi-infection OR multiinfection OR “simultaneous infection” OR “sequential infection “OR joint infection OR superinfection OR synergistic infection)

#### Web Of Science (307 Results)

(malaria OR Plasmodium OR Plasmodium falciparum OR “P. falciparum” OR “Plasmodium vivax" OR malarial parasite)
AND
(schistosom* OR schistosomiasis OR schistosoma OR schistosome OR bilharzia* OR Schistosoma mansoni OR Schistosoma haematobi* OR helminthiasis OR snail fever)
AND
(co-infection* OR "dual infection" OR "co-occurrence" OR cooccur* OR “co-distributed” OR “co-distribution” OR codistribut* OR "concomitant" OR polyparasitism OR multiparasitism OR multi-parasitism OR multi-infection OR multiinfection OR “simultaneous infection” OR “sequential infection “OR joint infection OR superinfection OR synergistic infection)

#### Global Index Medicus (3 Results)

(malaria OR Plasmodium OR Plasmodium falciparum OR “P. falciparum” OR “Plasmodium vivax" OR malarial parasite)
AND
(schistosom* OR schistosomiasis OR schistosoma OR schistosome OR bilharzia* OR Schistosoma mansoni OR Schistosoma haematobi* OR helminthiasis OR snail fever)
AND
(co-infection* OR "dual infection" OR "co-occurrence" OR cooccur* OR “co-distributed” OR “co-distribution” OR codistribut* OR "concomitant" OR polyparasitism OR multiparasitism OR multi-parasitism OR multi-infection OR multiinfection OR “simultaneous infection” OR “sequential infection “OR joint infection OR superinfection OR synergistic infection)

#### Pubmed (332 results)

(malaria OR Plasmodium OR Plasmodium falciparum OR “P. falciparum” OR “Plasmodium vivax" OR malarial parasite)
AND
(schistosom* OR schistosomiasis OR schistosoma OR schistosome OR bilharzia* OR Schistosoma mansoni OR Schistosoma haematobi* OR helminthiasis OR snail fever)
AND
(co-infection* OR "dual infection" OR "co-occurrence" OR cooccur* OR “co-distributed” OR “co-distribution” OR codistribut* OR "concomitant" OR polyparasitism OR multiparasitism OR multi-parasitism OR multi-infection OR multiinfection OR

“simultaneous infection” OR “sequential infection “OR joint infection OR superinfection OR synergistic infection)

#### S2 Text. Notes on data extraction

This table outlines whether the 2x2 contingency tables could be calculated for each study based on the data provided in the source papers.

| Study (Year) | Calculation Type | Possibility | Reason |
|---|---|---|---|
| **Duguay (2023)** | Sex vs. Co-infection | Not Possible | The breakdown of co-infection numbers by sex is not provided. |
| | Schisto (Any) vs. Malaria | Possible | - |
| **Sumbele (2021)** | Sex vs. Co-infection | Possible | - |
| | Schisto (haematobium) vs. Malaria | Possible | - |
| **Orish (2019)** | Sex vs. Co-infection | Not Possible | The sex of one child with a quadruple infection is not stated, preventing completion of the table. |
| | Schisto (haematobium) vs. Malaria | Possible | - |
| **Afolabi (2021)** | Sex vs. Co-infection | Possible | - |
| | Schisto (All Species) vs. Malaria | Possible | Data could be inferred by subtracting subgroups from totals provided in the paper. |
| **Getie (2015)** | Sex vs. Co-infection | Not Possible | Cannot be calculated because the underlying Schisto/Malaria table is not possible. |
| | Schisto (mansoni) vs. Malaria | Not Possible | The entire study population was malaria-positive, so there is no malaria-negative comparison group. |
| **Metafteria (2025)** | Sex vs. Co-infection | Not Possible | Cannot be calculated because the underlying Schisto/Malaria table is not possible. |
| | Schisto (mansoni) vs. Malaria | Not Possible | The entire study population was malaria-positive, so there |

| | | | |
|---|---|---|---|
| | | | is no malaria-negative comparison group. |
| **Florey (2012)** | Sex vs. Co-infection | Not Possible | The specific breakdown of how many co-infected individuals were male versus female is not provided. |
| | Schisto (haematobium) vs. Malaria | Possible | - |
| **Adedoja (2015)** | Sex vs. Co-infection | Possible | - |
| | Schisto (haematobium) vs. Malaria | Possible | - |
| | Schisto (mansoni) vs. Malaria | Not Possible | The specific number of children co-infected with both S. mansoni and P. falciparum is not provided. |
| **Oboh-Imafidon (2023)** | Sex vs. Co-infection | Not Possible | The breakdown of co-infection numbers by sex is not provided. |
| | Schisto (haematobium) vs. Malaria | Not Possible | The study population was composed entirely of S. haematobium-positive individuals, so there is no schisto-negative comparison group. |
| **Akosah-Brempong (2021)** | Sex vs. Co-infection | Possible | - |
| | Schisto (All Species) vs. Malaria | Possible | - |
| **Yapi (2015)** | Sex vs. Co-infection | Not Possible | The specific breakdown of co-infected children by sex is not provided. |
| | Schisto (Any) vs. Malaria | Possible | - |
| | Schisto (Species-specific) vs. Malaria | Not Possible | The paper does not provide specific co-infection numbers |

| | | | |
|---|---|---|---|
| | | | for S. mansoni and S. haematobium separately. |
| **Tine (2011)** | Sex vs. Co-infection | Not Possible | The breakdown of the 55 co-infected children by sex is not provided. |
| | Schisto (Any) vs. Malaria | Not Possible | The study was conducted exclusively among children who were already positive for malaria. |
| **Briand (2005)** | Sex vs. Co-infection & Schisto vs. Malaria | Not Possible | The study lacks a single point-in-time prevalence for malaria to cross-tabulate with schistosomiasis data, making a standard 2x2 table impossible. |
| **Dawaki (2019)** | Sex vs. Co-infection | Possible | - |
| | Schisto (mansoni & haematobium) vs. Malaria | Possible | - |
| | Schisto (Any) vs. Malaria | Not Possible | Combined schistosomiasis data also includes Soil-Transmitted Helminths and cannot be isolated. |
| **Dejon Agobe (2018)** | Sex vs. Co-infection | Not Possible | A breakdown of the co-infected group by sex is not specified. |
| | Schisto (haematobium) vs. Malaria | Possible | - |
| **Kamau (2021)** | Sex vs. Co-infection | Possible | - |
| | Schisto (Any/mansoni) vs. Malaria | Possible | The CCA test used is primarily sensitive for S. mansoni, so "Any" and "mansoni" are treated as the same. |
| **Mnkugwe (2020)** | Sex vs. Co-infection | Not Possible | The paper does not provide a breakdown of the co-infected group by sex. |
| | Schisto (Any) vs. Malaria | Possible | - |

| | | | |
|---|---|---|---|
| **Abay (2013)** | Sex vs. Co-infection | Not Possible | The underlying Schisto/Malaria table cannot be created as the entire study group was malaria-positive. |
| | Schisto (mansoni) vs. Malaria | Not Possible | The entire study population was malaria-positive, meaning there was no negative control group for comparison. |
| **Mulu (2013)** | Sex vs. Co-infection | Not Possible | The study does not provide a breakdown of co-infection cases by sex. |
| | Schisto (mansoni) vs. Malaria | Possible | Based on explicit numbers from Table 5, despite a data inconsistency in the text. |
| **Vengesai (2021)** | Sex vs. Co-infection | Not Possible | Data to populate the table is missing. |
| | Schisto (mansoni & haematobium) vs. Malaria | Possible | - |
| | Schisto (Any) vs. Malaria | Not Possible | The number of individuals co-infected with both S. mansoni and S. haematobium is not reported. |
| **Kamdem (2019)** | Sex vs. Co-infection | Possible | - |
| | Schisto (mansoni) vs. Malaria | Possible | - |
| **Kununghi (2017)** | Sex vs. Co-infection | Not Possible | The specific number of co-infected boys or girls is not provided. |
| | Schisto (mansoni) vs. Malaria | Possible | - |
| **Sokhna (2004)** | Sex vs. Co-infection | Not Possible | The total number of male and female participants in the cohort is not stated. |
| | Schisto (mansoni) vs. Malaria | Possible | - |
| **Doumbo (2014)** | Sex vs. Co-infection | Not Possible | The study does not provide a breakdown of the 39 co-infected individuals by sex. |

| | | | |
|---|---|---|---|
| | Schisto (haematobium) vs. Malaria | Possible | - |
| **Degarege (2012)** | Sex vs. Co-infection | Not Possible | The study does not provide a breakdown of co-infected individuals by sex. |
| | Schisto (mansoni) vs. Malaria | Possible | - |
| **Muhammed (2023)** | Sex vs. Co-infection | Possible | - |
| | Schisto (haematobium) vs. Malaria | Possible | - |
| **Lyke (2006)** | Sex vs. Co-infection | Possible | - |
| | Schisto vs. Malaria | Not Possible | The study has a prospective cohort design, not a cross-sectional one, making this calculation invalid. |
| **Makouloutou-Nzassi (2025)** | Sex vs. Co-infection | Not Possible | The study does not provide a sex breakdown for the 31 co-infected children. |
| | Schisto vs. Malaria | Not Possible | The malaria status of the schisto-negative children is not specified. |
| **Hurlimann (2019)** | Sex vs. Co-infection | Not Possible | The paper does not provide a breakdown of the co-infected groups by sex. |
| | Schisto (haematobium & mansoni) vs. Malaria | Possible | - |
| **Hürlimann (2014)** | Sex vs. Co-infection | Not Possible | The paper does not break down the number of co-infections by sex. |
| | Schisto (haematobium & mansoni) vs. Malaria | Possible | - |

| | | | |
|---|---|---|---|
| **Sangweme (2014)** | Sex vs. Co-infection | Not Possible | The study does not report the number of co-infected children stratified by sex. |
| | Schisto (Any) vs. Malaria | Possible | - |
| **Mene (2023)** | Sex vs. Co-infection | Not Possible | The number of individuals co-infected with schistosomiasis and malaria is not stratified by sex. |
| | Schisto (Any) vs. Malaria | Possible | - |

#### S3 Text. Quality appraisal tool

We used an adjusted version of the Quality Assessment Tool for Observational Cohort and Cross-Sectional Studies for Schistosomiasis based on [1] and National Institutes of Health (NIH, Bethesda, MD, United States of America) to perform the quality appraisal [2]. The adjusted tool for Malaria and Schistosomiasis was applied to all eligible study designs (cohort studies, cross-sectional studies, case-control studies, before-after studies, randomised controlled trials).

**Quality assessment categories**

| **Category** | **Question** | **Select 'yes' if** | **Select 'no' if** |
|---|---|---|---|
| Aim | Was the research question or objective in this paper clearly stated? | Authors clearly describe goal of their research and state the population, exposure and outcome, e.g. the aim was to identify risk factors of malaria and schistosomiasis co-infection in children | No aim or unspecific aim, e.g. this study examines the Malaria Schistosome co-infection patterns of an isolated farm-worker community |
| Representativeness | Was the study population clearly specified and defined? Were inclusion and exclusion criteria for being in the study prespecified and applied uniformly to all participants? (For case-control studies, were cases and | Authors describe the group of people from which the study participants were selected or recruited, using demographics, location, and time period; e.g. school-age children, male and female (aged 5-14), enrolled on Oct 1, 2014, in the five | Description lacking specifics on study population demographics, location, and time period, e.g. community members living in five communities were the study population |

| Category | Question | Select 'yes' if | Select ‘no’ if |
|---|---|---|---|
| | controls clearly differentiated?) | schools in the catchment areas of study villages were eligible | |
| Representativeness | Is the sampling method clearly described? Is the sample representative of the population from which it is drawn and was a sample size justification, power description, or variance and effect estimates provided? (For RCTs was the method of randomisation described and is risk of bias low? For case-control studies, was selection of cases and controls clearly described?) | Sampling method clearly described, e.g. ten school-age children (5-14) per class in each of the five schools were sampled using stratified random sampling, statistical power was assessed | Lacking details to understand how sampling was done, e.g. 50 adults per village were selected for inclusion in the study, no justification for sample size provided |

| Category | Question | Select 'yes' if | Select 'no' if |
|---|---|---|---|
| Representativeness | Response rate >50% or differences between respondents and non-respondents described? Was loss to follow-up after baseline 20% or less (if applicable)? | Provides statistics on response rate, e.g. 93% of respondents selected from village registries agreed to participate in the study, or differences in known characteristics of respondents and non-respondents (age, gender) were assessed using t-tests, loss to follow-up was 15% (if applicable) | Lacking information on response rate and differences between respondents and non-respondents provided |
| Exposure | Were risk factors for Malaria and Schistosome infection clearly defined and implemented consistently across all study participants? | **Yes:** Each shared risk factor (e.g., sociodemographic, environmental, spatial) has a clear definition provided in the study, and these definitions were applied consistently to all participants | No: Risk factor definitions are unclear, missing, or inconsistently applied across participants. |
| Outcomes | Were malaria and schistosomiasis diagnoses based on valid and reliable methods? | Diagnosis of malaria and schistosomiasis is confirmed through microscopy, antigen tests, or antibody tests. | The diagnostic methods are based on self-report, are not clearly described. |

| Category | Question | Select 'yes' if | Select ‘no’ if |
|---|---|---|---|
| Outcomes | Was the definition of co-infection clearly stated and was the reference group clearly defined? | The study clearly defines co-infection, including whether it is concurrent or sequential and explicitly states the reference group. | The definition of co-infection and the resulting reference group is absent or ambiguous. |
| Outcomes | Were exposures and outcomes measured independently by two people? | Measurement and diagnostic test done by different people, e.g. self-reported water contact using survey (exposure) and microscopy by a trained technician (outcome) | Same person administering questions on water contact, mosquito exposure and infection, e.g. technicians responsible for microscopy (outcome measurement) recruited to contact direct water contact observation (exposure) |

| Category | Question | Select 'yes' if | Select 'no' if |
|---|---|---|---|
| Analysis | Were adjusted effect estimates shown? For example, were age and gender adjusted for where relevant? | Studies report both types of models, e.g. unadjusted association between risk factors and infection status . Confidence Intervals are provided | Studies report only adjusted or unadjusted ORs/RRs, fail to provide 95% CIs |
